# Analysis of Changes in the Burden of Nutritional Deficiencies in China, the G20, and Globally from 1990 to 2021 Based on the Global Burden of Disease 2021 Data

**DOI:** 10.1101/2025.04.24.25326366

**Authors:** Lin Ma, Xiangwen Li, Xufei Zhao, Ruonan Zhao, Rongrong Chen, Rongqiang Zhang

## Abstract

**Objective:** Nutritional deficiency, has recently received considerable attention as significant risk factor affecting global health. Nutritional deficiencies are a significant global burden associated with chronic disease. This study analyzed the burden of nutritional deficiencies in China from 1990 to 2021 based on age and gender, and compared it with the burden observed in global and G20 regions. This study aimed to understand the trends in nutritional deficiencies in China using this burden analysis and evaluate the changes in their burden to guide future public health policies.

**Method:** This study utilized date from the global burden of disease database (GBD) for 2021, on the prevalence and years lived with disability (YLDs) attributed to nutritional deficiencies in China from 1990 to 2021. We analyzed the burden and temporal trends of nutritional deficiency in China based on gender, age, and region. Additionally, we analyzed the disease burden in the global and G20 regions, and compared it with the Chinese disease burden. We used estimated annual percentage change to reflect disease trends and subsequently utilized relevant cutting-edge technologies to predict the trends of nutritional deficiencies in China and global regions for 2035.

**Result:** The burden of nutritional deficiencies in China demonstrated a significant declining trend from 1990 to 2021. China had more male patients with nutritional deficiencies in the < 5 age group than female patients, while the proportion of female patients was higher than that of males in other age groups, especially during pregnancy. Furthermore, dietary iron deficiency accounted for most of nutritional deficiency cases, contributing the largest share of the burden. From 1990 to 2021, epidemiological changes were the primary driver of the increased disease burden of nutritional deficiencies in China.

**Conclusion:** The disease burden of nutritional deficiencies in China demonstrated a decreasing trend from 1990 to 2021. Meanwhile, the disease burden of nutritional deficiencies in the G20 and global regions demonstrated a significant declining trend. This may be associated with socioeconomic advancement. Regarding gender and age, the number of cases of nutritional deficiencies is highest among individuals aged 30 to 34 in China, with the highest number of female patients for nutritional deficiency far exceeding that of male patients of the same age group. This is associated with social factors, including poverty and social inequality. Social inequality and poverty increase the risk of illness among women. Regarding diseases, the disease burden of dietary iron deficiency is relatively high, followed by vitamin A deficiency. This may be linked to regional differences and dietary patterns in China.

## 1 Introduction

Nutrition provides the energy required for all processes in the human body. In China, nutritional deficiencies are defined as disease symptoms caused by insufficient nutrient intake, including vitamins, proteins, and trace elements. Nutritional deficiency is defined as a severe reduction in one or more nutrient levels, that prevents the body from functioning normally and increases the risk of multiple diseases, including cancer, diabetes, and heart disease^[1]^. Nutritional deficiency includes protein-energy malnutrition (PEM), iron deficiency anemia, iodine, vitamin A deficiency and other nutritional deficiencies^[2]^.

Although there is no consensus on the definition and clinical evaluation of NDs, they usually include PEM and micronutrient deficiencies^[3]^. PEM is a series of diseases resulting from deficiencies in all macronutrients, including an intermediate state between malnutrition and severe malnutrition (cachexia). PEM is a common nutritional issue globally, evident in developed and developing countries^[4]^.Additionally, PEM can compromise immune responses, endangering public health, especially in children and the elderly, potentially resulting in death^[5]^.Despite the decreasing incidence of PEM in children due to improvements in healthcare and food industry advancement, it continues to present a significant health burden on people of all age groups. Addressing this public health issue is our primary responsibility^[6]^.Micronutrient deficiencies, including iron, vitamin A, and iodine deficiencies, result in anemia, night blindness, and goiter, respectively, which jeopardize the health of > 2 billion people globally^[7]^.Vitamin A deficiency can result in various eye diseases, including xerophthalmia, ranging from night blindness to more severe clinical conditions such as corneal softening, scarring and permanent blindness^[8]^. Vitamin A deficiency is associated with an increased risk of measles and diarrhea-related deaths in children^[9]^.Vitamin A deficiency can lead to various clinical outcomes, including night blindness. Measles is another disease caused by vitamin A deficiency^[10]^.Management of vitamin A deficiency begins with food, including supplementing foods rich in vitamin A (animal products) or increasing the intake of β-carotene-rich diets, which is a more sustainable solution. However, high costs, accessibility issues, and cultural dietary habits limit its potential to alleviate vitamin A deficiency. This r issue that needs requires resolution^[11]^.

Iron deficiency is a leading risk factor for disability and death, affecting approximately 2 billion people. Nutritional iron deficiency occurs when dietary iron absorption fails to meet physiological requirements. Iron deficiency is the most common micronutrient deficiency and cause of anemia globally; approximately 42% of anemia cases in children <5 years old are attributable to iron deficiency^[12][13]^. Iron deficiency anemia may result in immune system dysfunction, gastrointestinal disorders, and impaired thermoregulation and neurocognitive function^[14]^. In addition, anemia may be a risk or prognostic factor for other diseases (tuberculosis and heart failure)^[15][16]^. Anemia remains a major health problem, particularly among women in underdeveloped countries, necessitating the implementation of prevention programs aimed at improving access to iron supplements, early diagnosis, and hemoglobinopathy treatment^[17]^. Iodine deficiency contributes to a global disease burden, impairing thyroid hormone synthesis and resulting in various metabolic and growth-related diseases, thereby jeopardizing physical health and development^[18][19]^. The synthesis of thyroid hormone depends on iodine intake, an important precursor for thyroid hormone production. Thyroid hormone is essential for growth and development^[20]^. Thyroid hormone deficiency in early childhood can result in irreversible neurological developmental disorders known as cretinism^[21]^. Kittens disease is characterized by cognitive and growth retardation and incomplete tooth development. Individuals with intellectual disabilities are more prone to encountering difficulties in accessing equitable health care and experience higher rates of premature death compared to the general population^[22]^.

The G20 is an international economic cooperation forum comprising 19 countries and the European Union. The members include Argentina, Australia, Brazil, Canada, China, France, Germany, India, Indonesia, Italy, Japan, South Korea, Mexico, Russia, Saudi Arabia, South Africa, Turkey, the United Kingdom, and the United States. The G20 was established to promote international financial stability, foster global economic cooperation, and address global economic issues. Established in 1999, the G20 was initially composed of finance ministers and central bank governors from various countries. Following the 2008 financial crisis, the G20 leadership summit mechanism was formally established, making it the primary platform for global economic governance. The G20 member countries represent approximately 85% of the global GDP, >75% of global trade, and approximately 66.67% of the global population. The health burden on the populations of these G20 countries is closely associated with their specific economic environment and their medical and health conditions^[23]^.

The global burden of disease (GBD), injury and risk factors is an ongoing extensive research initiative that describes global health status by estimating essential global, regional and national health indicators. The latest research on the status and trends of G20 in the GBD 2021 study offers direction for formulating clinical care guidelines and public health initiatives, which assist policymakers in the precise and effective allocation of healthcare resources, thereby alleviating social and individual burdens^[24]^. Therefore, this study utilized data on malnutrition prevalence and years of lost disability life from the GBD 2021 database, categorized by gender, age, and time. This study aimed to analyze the disease burden and trends of nutritional deficiencies in China.

## 2 Data sources and methods

### 2.1 Data sources

The data for this study were obtained from the GBD 2021 database within the GBD study, which encompasses nutritional deficiencies, including PEM, iron deficiency anemia, iodine, vitamin A, and other nutrient deficiencies. The GBD 2021 Disease and Injury Burden Analysis utilized 100,983 data sources to estimate years lived with disability (YLD), years of lost life (YLL), disability-adjusted life years (DALY), and healthy life expectancy (HALE) for 371 diseases and injuries. Data were obtained from life registration systems, verbal autopsies, censuses, household surveys, specialized disease registers, health service linkage data, and other sources. The YLD calculation method for each disease and injury involves multiplying the prevalence of the condition by its respective disability weight for the year of onset, adjusted for age, sex, location, and specific year. This study must guarantee that no patient privacy or personal identity disclosure data were utilized. All relevant legal and ethical guidelines were followed while utilizing GBD data, and knowledge sharing was executed under a non-commercial use license^[25]^.The study analyzed data from the GBD2021 which offers the latest estimates of epidemiological data regarding the burden of 371 diseases and injuries across 21 GBD regions and 204 countries and territories from 1990 to 2021. These data are available through the Global Health Data Exchange (https://ghdx.healthdata.org/gbd-2021/sources) ^[26]^.

### 2.2 Method

Common research surveys employ four leading indicators to evaluate the disease burden: incidence, prevalence, YLL and DALYs. The DALYs, an indicator used to measure disease burden, include YLL due to premature death and YLL due to YLD. To adjust for differences in age structure across different populations, the ratios of these indicators are calculated by dividing each corresponding indicator by the population size^[27]^. Herein, we primarily utilize YLDs in epidemiology, denoting YLD, which is years of life lost due to disability. It is an indicator used to measure the loss of healthy life years attributed to diseases or poor health status. It is calculated by multiplying the prevalence of a disease by its associated disability weight. This indicator elucidates impact of diseases on the quality of life and is an important component of GBD research. This study employed prevalence and YLDs as primary indicators to illustrate the disease burden and trends of nutritional deficiencies in China.

The Estimated Annual Percentage Change (EAPC) is commonly utilized metric for assessing rate trends over specific time intervals. It is calculated using a linear regression model represented by the equation Y = α + βX + ε, where X represents the calendar year, Y represents the natural logarithm of each age-standardized indicator, and ε represents the error term. The EAPC is calculated as 100 × (exp(β) -1). The corresponding 95% confidence interval (CI) for these estimates is calculated using the previously mentioned linear regression model. A 95% CI < 0 indicates a declining trend, while a CI > 0 indicates an increasing trend^[27]^.

Prediction: We used the Bayesian age-period cohort model (BAPC) to predict the future trend of nutritional deficiencies in China and globally.

Average annual percentage change (AAPC) describes the average annual change rate of a disease metric including incidence and mortality) over a specified timeframe. It provides a more comprehensive trend analysis by integrating data from several years, rather than simply comparing the data from the first and last two years. The calculation of AAPC involves the following steps: Estimating the joinpoint model: Initially, we used joinpoint software or alternative statistical methods to estimate the best-fit joinpoint model for the data. The joinpoint model uses segmented linear regression to describe the trend changes in the data across different periods. Calculating the annual percentage change (APC) for each interval: For each segmented interval, we calculated its APC. AAPC is calculated by weighing the APCs of each interval based on their lengths. AAPC provides a single number that describes the average change trend over various years, even if the trend has changed during this period. This method allows researchers to describe the overall trend change with a single indicator, even when there are multiple trend change points^[28]^.

### 2.3 Statistical analysis

The indicators analyzed in this study for prevalence and YLDs primarily include quantity, ratio, and percentage. The calculation method for YLDs is to multiply the age-specific prevalence of nutritional deficiencies by their corresponding disability weights. The calculation method for YLLs involves multiplying age-specific mortality rates by the standard life expectancy at the age of death. DALY is calculated by summing YLDs and YLLs. At this stage, each model was executed until convergence, after which the 95^th^ and 25^th^ ordered samples from 1,000 posterior models were obtained as the 95% uncertainty interval (UI) for each point estimate^[27]^. We used Excel to structure the data and subsequently utilized Statistical Package for the Social Sciences software to verify that the conditions for linear regression were met. R software was used for all statistical analyses. A *P*-values < 0.05 was considered statistically significant.

## 3 Results

### 3.1 Analysis of the disease burden of nutritional deficiencies in China, the global region, and the G20 region from 1990 to 2021

Table 1 indicates that the global YLDs attributed to nutritional deficiency in 2021 were approximately 38.64 million cases (95% UI: 21.63– 55.25 million). The age-standardized rate of YLDs was 507.94 cases (95% UI: 343.56–726.80) per 100,000 population. The 2021 global YLDs attributed to nutritional deficiency were approximately 36.78 million cases (95% UI: 25.03–51.75 million), and the age-standardized rate of YLDs was 664.12 cases (95% UI: 453.17–933.22) per 100,000 population compared to those of 1990. EAPC exhibited an average annual decrease of 0.86% (95% CI: –0.92% to –0.79%) (Table 1).

**Table 1.**
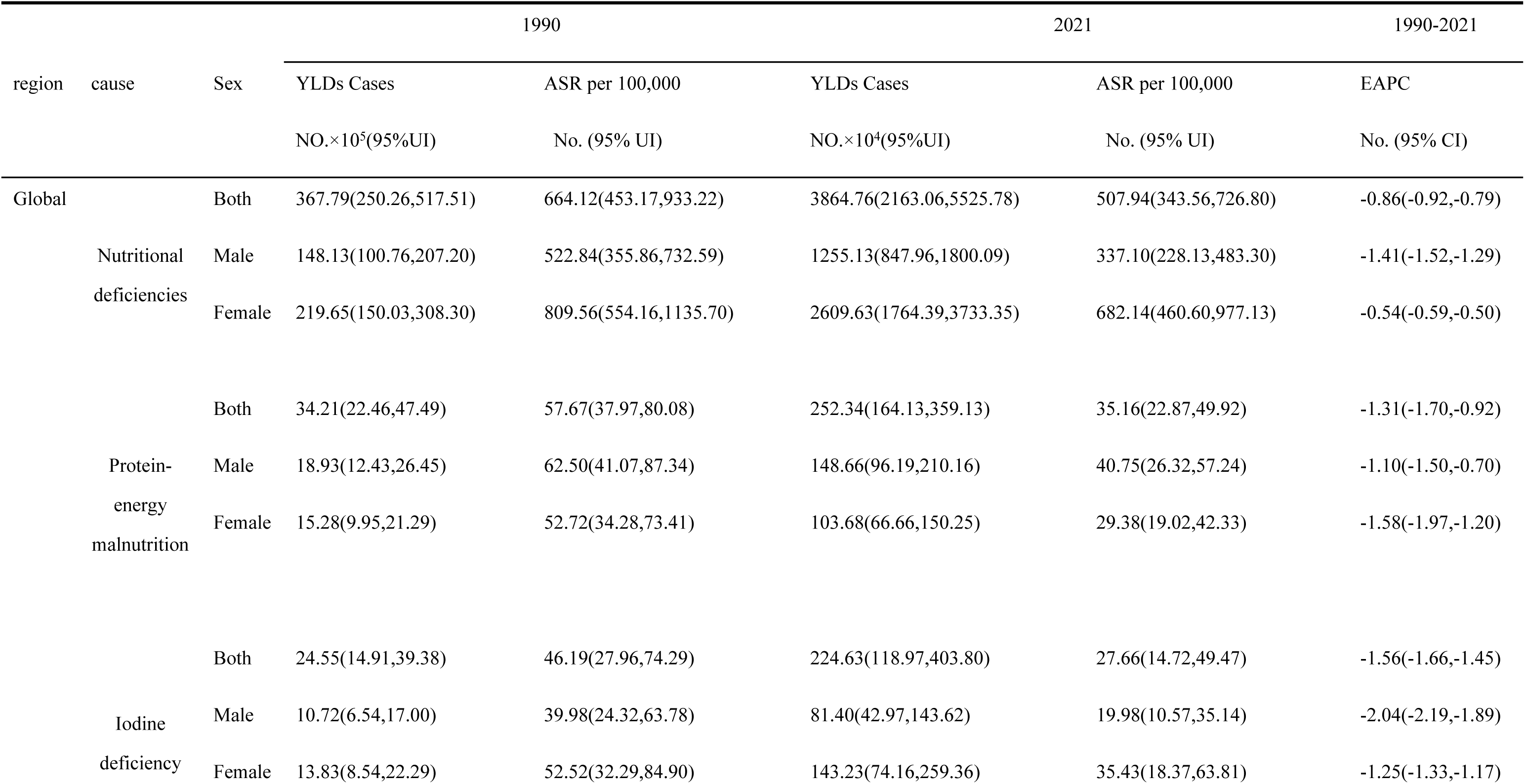
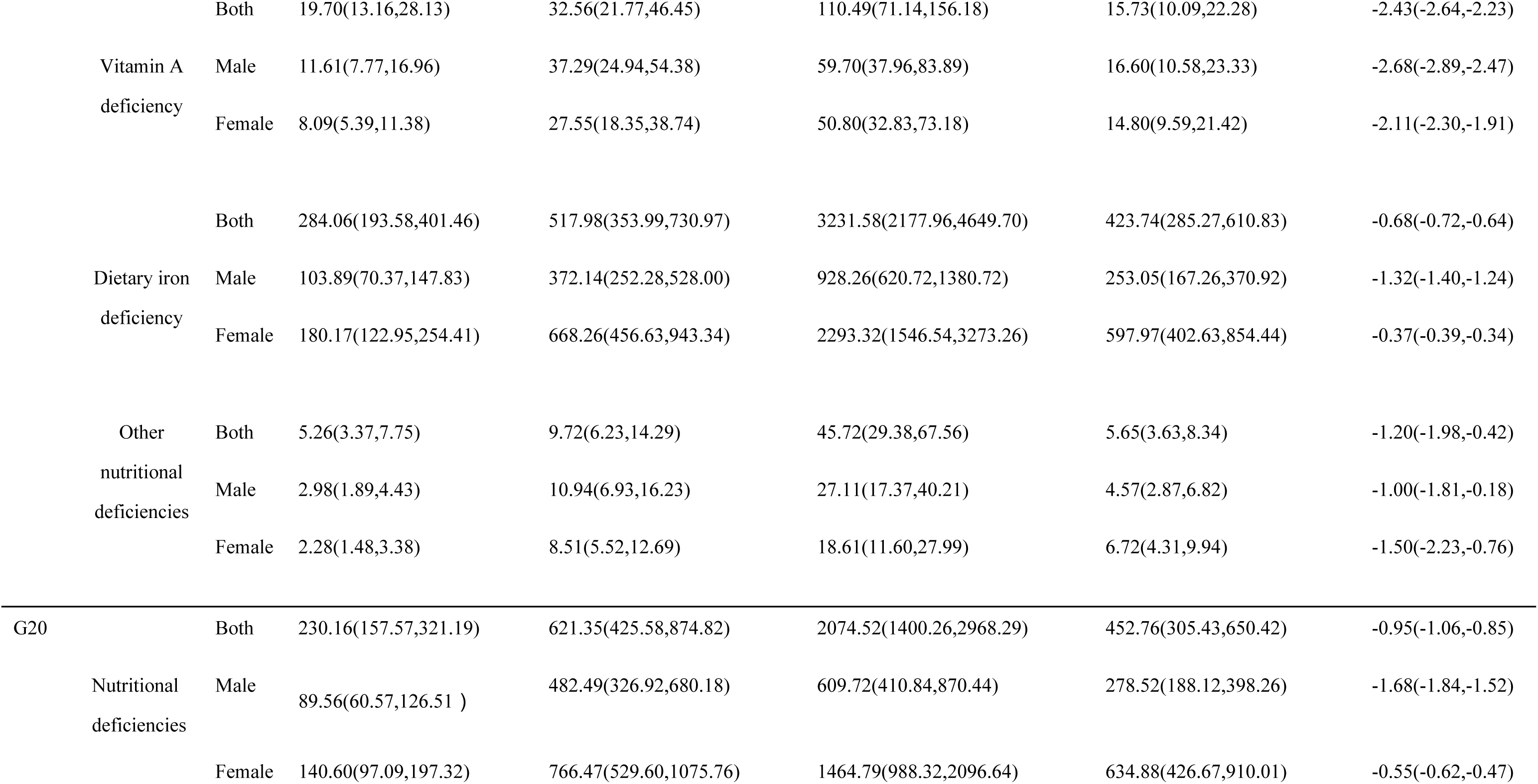
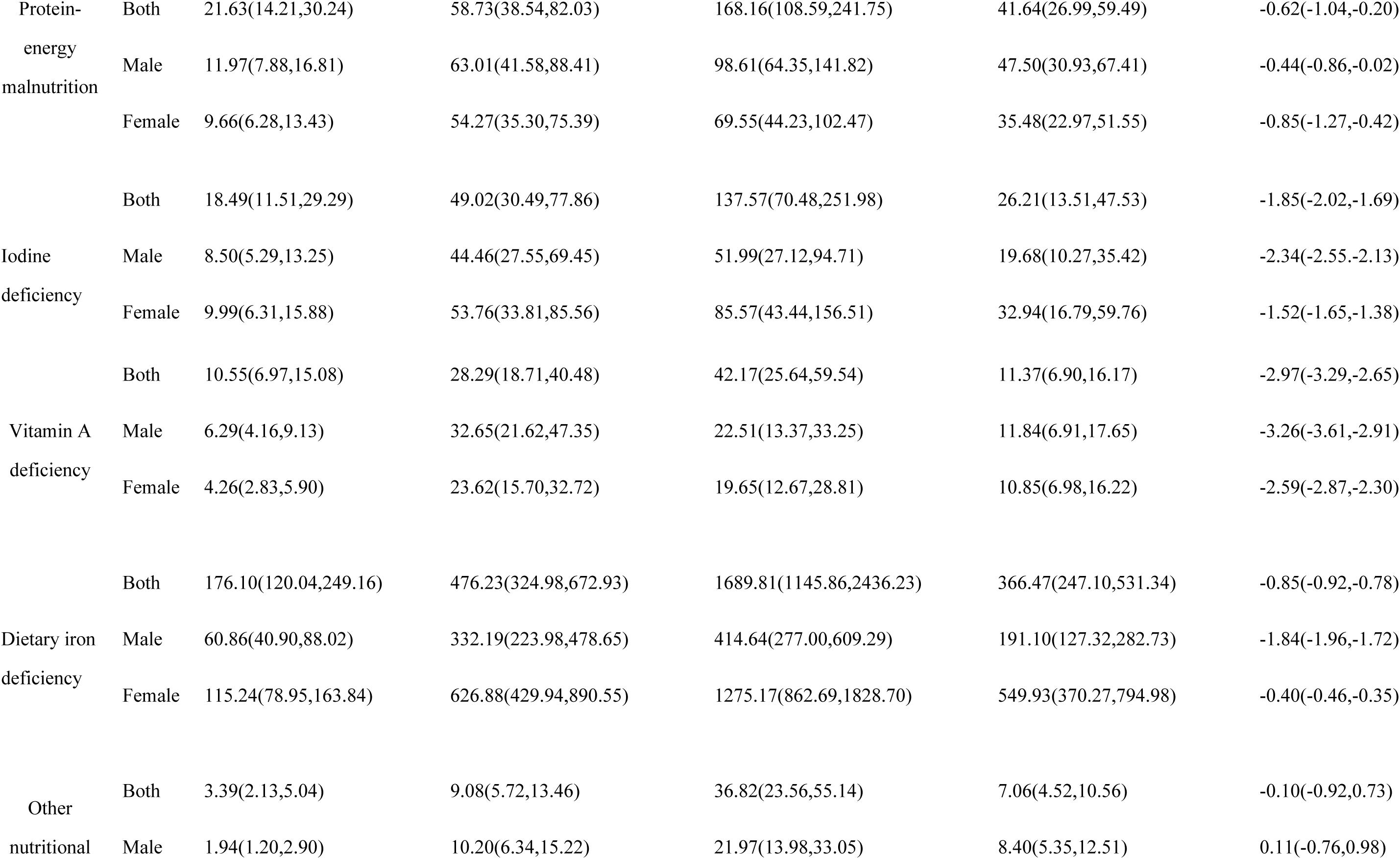
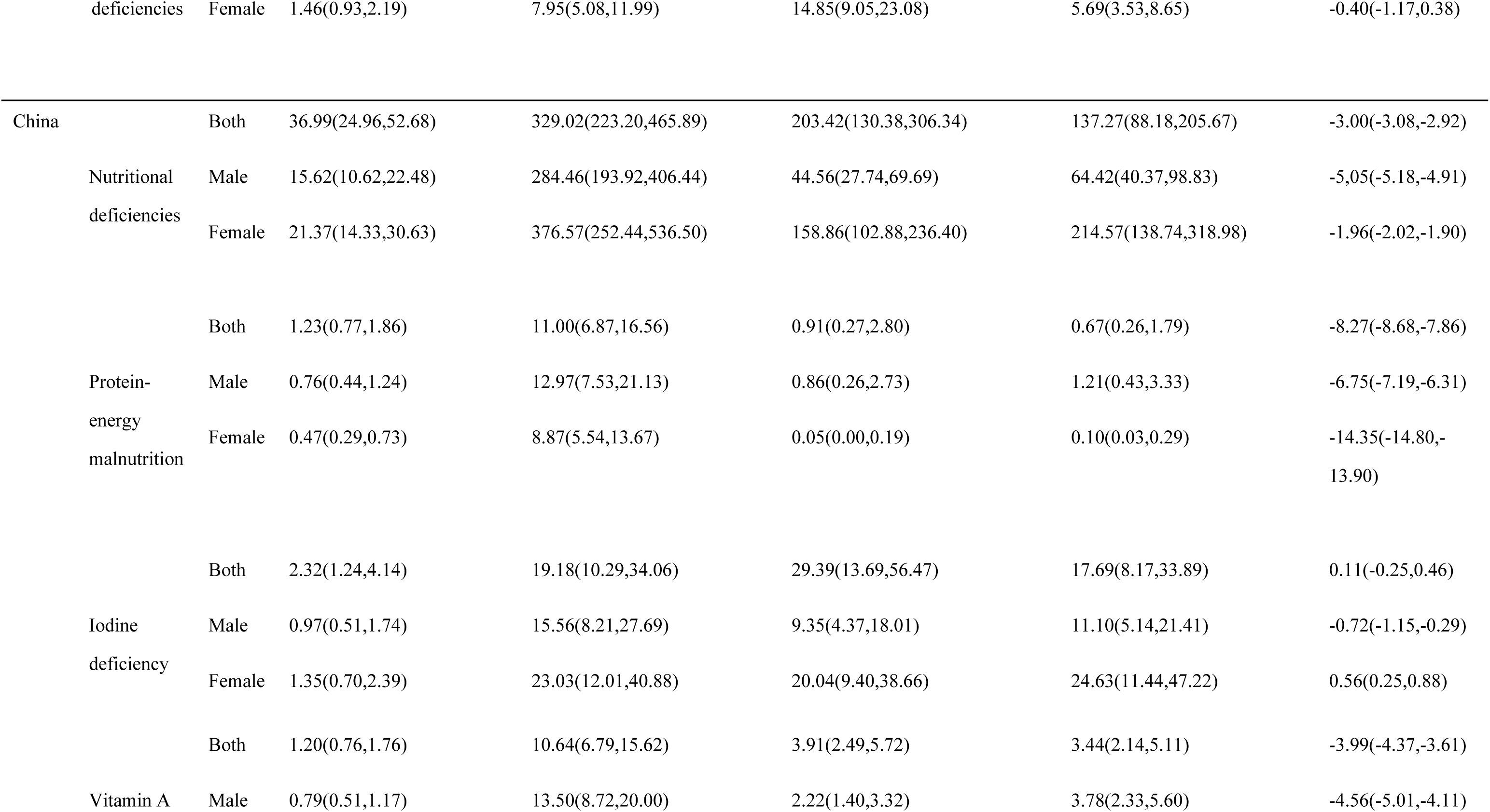
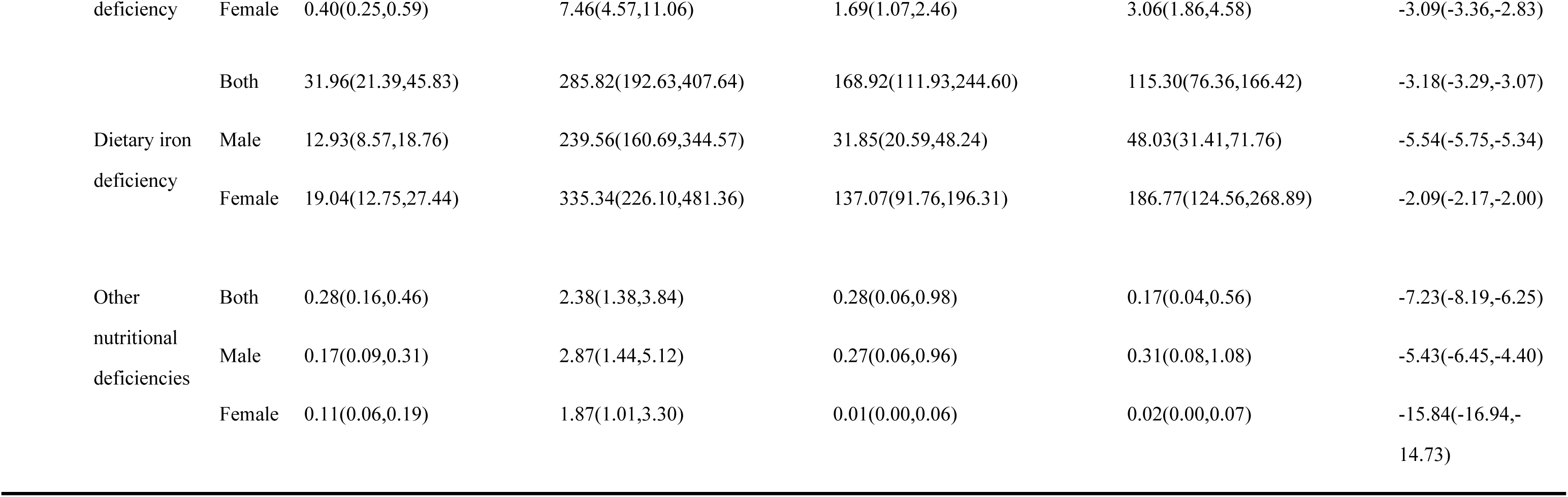
The YLDs levels of nutritional deficiency in China, the G20, and Globally from 1990 to 2021.

| region | cause | Sex | 1990 |  | 2021 |  | 1990-2021 |
| --- | --- | --- | --- | --- | --- | --- | --- |
|  |  |  | YLDs Cases | ASR per 100,000 | YLDs Cases | ASR per 100,000 | EAPC |
| | | | NO.$\times 10^5$(95%UI) | No. (95% UI) | NO.$\times 10^4$(95%UI) | No. (95% UI) | No. (95% CI) |
| Global | Nutritional deficiencies | Both | 367.79(250.26,517.51) | 664.12(453.17,933.22) | 3864.76(2163.06,5525.78) | 507.94(343.56,726.80) | -0.86(-0.92,-0.79) |
|  |  | Male | 148.13(100.76,207.20) | 522.84(355.86,732.59) | 1255.13(847.96,1800.09) | 337.10(228.13,483.30) | -1.41(-1.52,-1.29) |
|  |  | Female | 219.65(150.03,308.30) | 809.56(554.16,1135.70) | 2609.63(1764.39,3733.35) | 682.14(460.60,977.13) | -0.54(-0.59,-0.50) |
|  | Protein-energy malnutrition | Both | 34.21(22.46,47.49) | 57.67(37.97,80.08) | 252.34(164.13,359.13) | 35.16(22.87,49.92) | -1.31(-1.70,-0.92) |
|  |  | Male | 18.93(12.43,26.45) | 62.50(41.07,87.34) | 148.66(96.19,210.16) | 40.75(26.32,57.24) | -1.10(-1.50,-0.70) |
|  |  | Female | 15.28(9.95,21.29) | 52.72(34.28,73.41) | 103.68(66.66,150.25) | 29.38(19.02,42.33) | -1.58(-1.97,-1.20) |
|  | Iodine deficiency | Both | 24.55(14.91,39.38) | 46.19(27.96,74.29) | 224.63(118.97,403.80) | 27.66(14.72,49.47) | -1.56(-1.66,-1.45) |
|  |  | Male | 10.72(6.54,17.00) | 39.98(24.32,63.78) | 81.40(42.97,143.62) | 19.98(10.57,35.14) | -2.04(-2.19,-1.89) |
|  |  | Female | 13.83(8.54,22.29) | 52.52(32.29,84.90) | 143.23(74.16,259.36) | 35.43(18.37,63.81) | -1.25(-1.33,-1.17) |

|  |  |  |  |  |  |  |
| --- | --- | --- | --- | --- | --- | --- |
|  | Both | 19.70(13.16,28.13) | 32.56(21.77,46.45) | 110.49(71.14,156.18) | 15.73(10.09,22.28) | -2.43(-2.64,-2.23) |
| Vitamin A deficiency | Male | 11.61(7.77,16.96) | 37.29(24.94,54.38) | 59.70(37.96,83.89) | 16.60(10.58,23.33) | -2.68(-2.89,-2.47) |
|  | Female | 8.09(5.39,11.38) | 27.55(18.35,38.74) | 50.80(32.83,73.18) | 14.80(9.59,21.42) | -2.11(-2.30,-1.91) |
|  | Both | 284.06(193.58,401.46) | 517.98(353.99,730.97) | 3231.58(2177.96,4649.70) | 423.74(285.27,610.83) | -0.68(-0.72,-0.64) |
| Dietary iron deficiency | Male | 103.89(70.37,147.83) | 372.14(252.28,528.00) | 928.26(620.72,1380.72) | 253.05(167.26,370.92) | -1.32(-1.40,-1.24) |
|  | Female | 180.17(122.95,254.41) | 668.26(456.63,943.34) | 2293.32(1546.54,3273.26) | 597.97(402.63,854.44) | -0.37(-0.39,-0.34) |
| Other nutritional deficiencies | Both | 5.26(3.37,7.75) | 9.72(6.23,14.29) | 45.72(29.38,67.56) | 5.65(3.63,8.34) | -1.20(-1.98,-0.42) |
|  | Male | 2.98(1.89,4.43) | 10.94(6.93,16.23) | 27.11(17.37,40.21) | 4.57(2.87,6.82) | -1.00(-1.81,-0.18) |
|  | Female | 2.28(1.48,3.38) | 8.51(5.52,12.69) | 18.61(11.60,27.99) | 6.72(4.31,9.94) | -1.50(-2.23,-0.76) |
| G20 | Both | 230.16(157.57,321.19) | 621.35(425.58,874.82) | 2074.52(1400.26,2968.29) | 452.76(305.43,650.42) | -0.95(-1.06,-0.85) |
| Nutritional deficiencies | Male | 89.56(60.57,126.51 ) | 482.49(326.92,680.18) | 609.72(410.84,870.44) | 278.52(188.12,398.26) | -1.68(-1.84,-1.52) |
|  | Female | 140.60(97.09,197.32) | 766.47(529.60,1075.76) | 1464.79(988.32,2096.64) | 634.88(426.67,910.01) | -0.55(-0.62,-0.47) |
| Protein-energy malnutrition | Both | 21.63(14.21,30.24) | 58.73(38.54,82.03) | 168.16(108.59,241.75) | 41.64(26.99,59.49) | -0.62(-1.04,-0.20) |
|  | Male | 11.97(7.88,16.81) | 63.01(41.58,88.41) | 98.61(64.35,141.82) | 47.50(30.93,67.41) | -0.44(-0.86,-0.02) |
|  | Female | 9.66(6.28,13.43) | 54.27(35.30,75.39) | 69.55(44.23,102.47) | 35.48(22.97,51.55) | -0.85(-1.27,-0.42) |
| Iodine deficiency | Both | 18.49(11.51,29.29) | 49.02(30.49,77.86) | 137.57(70.48,251.98) | 26.21(13.51,47.53) | -1.85(-2.02,-1.69) |
|  | Male | 8.50(5.29,13.25) | 44.46(27.55,69.45) | 51.99(27.12,94.71) | 19.68(10.27,35.42) | -2.34(-2.55,-2.13) |
|  | Female | 9.99(6.31,15.88) | 53.76(33.81,85.56) | 85.57(43.44,156.51) | 32.94(16.79,59.76) | -1.52(-1.65,-1.38) |
| Vitamin A deficiency | Both | 10.55(6.97,15.08) | 28.29(18.71,40.48) | 42.17(25.64,59.54) | 11.37(6.90,16.17) | -2.97(-3.29,-2.65) |
|  | Male | 6.29(4.16,9.13) | 32.65(21.62,47.35) | 22.51(13.37,33.25) | 11.84(6.91,17.65) | -3.26(-3.61,-2.91) |
|  | Female | 4.26(2.83,5.90) | 23.62(15.70,32.72) | 19.65(12.67,28.81) | 10.85(6.98,16.22) | -2.59(-2.87,-2.30) |
| Dietary iron deficiency | Both | 176.10(120.04,249.16) | 476.23(324.98,672.93) | 1689.81(1145.86,2436.23) | 366.47(247.10,531.34) | -0.85(-0.92,-0.78) |
|  | Male | 60.86(40.90,88.02) | 332.19(223.98,478.65) | 414.64(277.00,609.29) | 191.10(127.32,282.73) | -1.84(-1.96,-1.72) |
|  | Female | 115.24(78.95,163.84) | 626.88(429.94,890.55) | 1275.17(862.69,1828.70) | 549.93(370.27,794.98) | -0.40(-0.46,-0.35) |
| Other nutritional | Both | 3.39(2.13,5.04) | 9.08(5.72,13.46) | 36.82(23.56,55.14) | 7.06(4.52,10.56) | -0.10(-0.92,0.73) |
|  | Male | 1.94(1.20,2.90) | 10.20(6.34,15.22) | 21.97(13.98,33.05) | 8.40(5.35,12.51) | 0.11(-0.76,0.98) |
|  | deficiencies | Female | 1.46(0.93,2.19) | 7.95(5.08,11.99) | 14.85(9.05,23.08) | 5.69(3.53,8.65) | -0.40(-1.17,0.38) |
| China |  | Both | 36.99(24.96,52.68) | 329.02(223.20,465.89) | 203.42(130.38,306.34) | 137.27(88.18,205.67) | -3.00(-3.08,-2.92) |
|  | Nutritional deficiencies | Male | 15.62(10.62,22.48) | 284.46(193.92,406.44) | 44.56(27.74,69.69) | 64.42(40.37,98.83) | -5.05(-5.18,-4.91) |
|  |  | Female | 21.37(14.33,30.63) | 376.57(252.44,536.50) | 158.86(102.88,236.40) | 214.57(138.74,318.98) | -1.96(-2.02,-1.90) |
|  |  | Both | 1.23(0.77,1.86) | 11.00(6.87,16.56) | 0.91(0.27,2.80) | 0.67(0.26,1.79) | -8.27(-8.68,-7.86) |
|  | Protein-energy malnutrition | Male | 0.76(0.44,1.24) | 12.97(7.53,21.13) | 0.86(0.26,2.73) | 1.21(0.43,3.33) | -6.75(-7.19,-6.31) |
|  |  | Female | 0.47(0.29,0.73) | 8.87(5.54,13.67) | 0.05(0.00,0.19) | 0.10(0.03,0.29) | -14.35(-14.80,-13.90) |
|  |  | Both | 2.32(1.24,4.14) | 19.18(10.29,34.06) | 29.39(13.69,56.47) | 17.69(8.17,33.89) | 0.11(-0.25,0.46) |
|  | Iodine deficiency | Male | 0.97(0.51,1.74) | 15.56(8.21,27.69) | 9.35(4.37,18.01) | 11.10(5.14,21.41) | -0.72(-1.15,-0.29) |
|  |  | Female | 1.35(0.70,2.39) | 23.03(12.01,40.88) | 20.04(9.40,38.66) | 24.63(11.44,47.22) | 0.56(0.25,0.88) |
|  |  | Both | 1.20(0.76,1.76) | 10.64(6.79,15.62) | 3.91(2.49,5.72) | 3.44(2.14,5.11) | -3.99(-4.37,-3.61) |
|  | Vitamin A | Male | 0.79(0.51,1.17) | 13.50(8.72,20.00) | 2.22(1.40,3.32) | 3.78(2.33,5.60) | -4.56(-5.01,-4.11) |

|  |  |  |  |  |  |  |
| --- | --- | --- | --- | --- | --- | --- |
| deficiency | Female | 0.40(0.25,0.59) | 7.46(4.57,11.06) | 1.69(1.07,2.46) | 3.06(1.86,4.58) | -3.09(-3.36,-2.83) |
|  | Both | 31.96(21.39,45.83) | 285.82(192.63,407.64) | 168.92(111.93,244.60) | 115.30(76.36,166.42) | -3.18(-3.29,-3.07) |
| Dietary iron | Male | 12.93(8.57,18.76) | 239.56(160.69,344.57) | 31.85(20.59,48.24) | 48.03(31.41,71.76) | -5.54(-5.75,-5.34) |
| deficiency | Female | 19.04(12.75,27.44) | 335.34(226.10,481.36) | 137.07(91.76,196.31) | 186.77(124.56,268.89) | -2.09(-2.17,-2.00) |
| Other | Both | 0.28(0.16,0.46) | 2.38(1.38,3.84) | 0.28(0.06,0.98) | 0.17(0.04,0.56) | -7.23(-8.19,-6.25) |
| nutritional | Male | 0.17(0.09,0.31) | 2.87(1.44,5.12) | 0.27(0.06,0.96) | 0.31(0.08,1.08) | -5.43(-6.45,-4.40) |
| deficiencies | Female | 0.11(0.06,0.19) | 1.87(1.01,3.30) | 0.01(0.00,0.06) | 0.02(0.00,0.07) | -15.84(-16.94,-<br>14.73) |

In 2021, the G20 group had approximately 20.74 million cases (95% UI:14.00 million to 29.68 million), compared with 1990, the YLDs of G20 group attributed to nutritional deficiency were approximately 23.02 million cases (95% UI:15.76 million to 32.12 million), with the age-standardized rate of YLDs, there were 452.76 cases per 100,000 population (95% UI:305.43 to 650.42) in 2021 compared to 621.35 cases per 100,000 population in 1990 (95% UI: 425.58 to 874.82). The EAPC has an average annual decrease of 0.95% (95% CI: -1.06% to -0.85%).

In 2021, China had approximately 2.03 million cases (95% UI: 1.37–2.05 million), while the YLDs of China attributed to nutritional deficiency were approximately 3.69 million cases (95% UI: 2.49–5.27 million) in 1990. The age-standardized rate of YLDs was 137.27 per 100,000 population (95% UI: 88.18–205.67). In 1990, the rate was 329.02 per 100,000 population (95% UI: 223.20–465.89). The EAPC exhibited an average annual decrease of 3.00% (95% CI: –3.08% to –2.92%).

Table 2 indicates the prevalence attributed to nutritional deficiency by global region in 2021 was approximately 1.84 billion (95% UI: 1.81– 1.88 billion), with an ASPR of 23858.99 per 100,000 population (95% UI: 23445.77–24320.82) compared to 32217.95 per 100,000 population (95% UI: 31693.55–32740.92) in 1990. The ASPR EAPC exhibited a decreasing trend of 0.98% annually (95% CI: –1.00% to –0.96%).

**Table 2.**
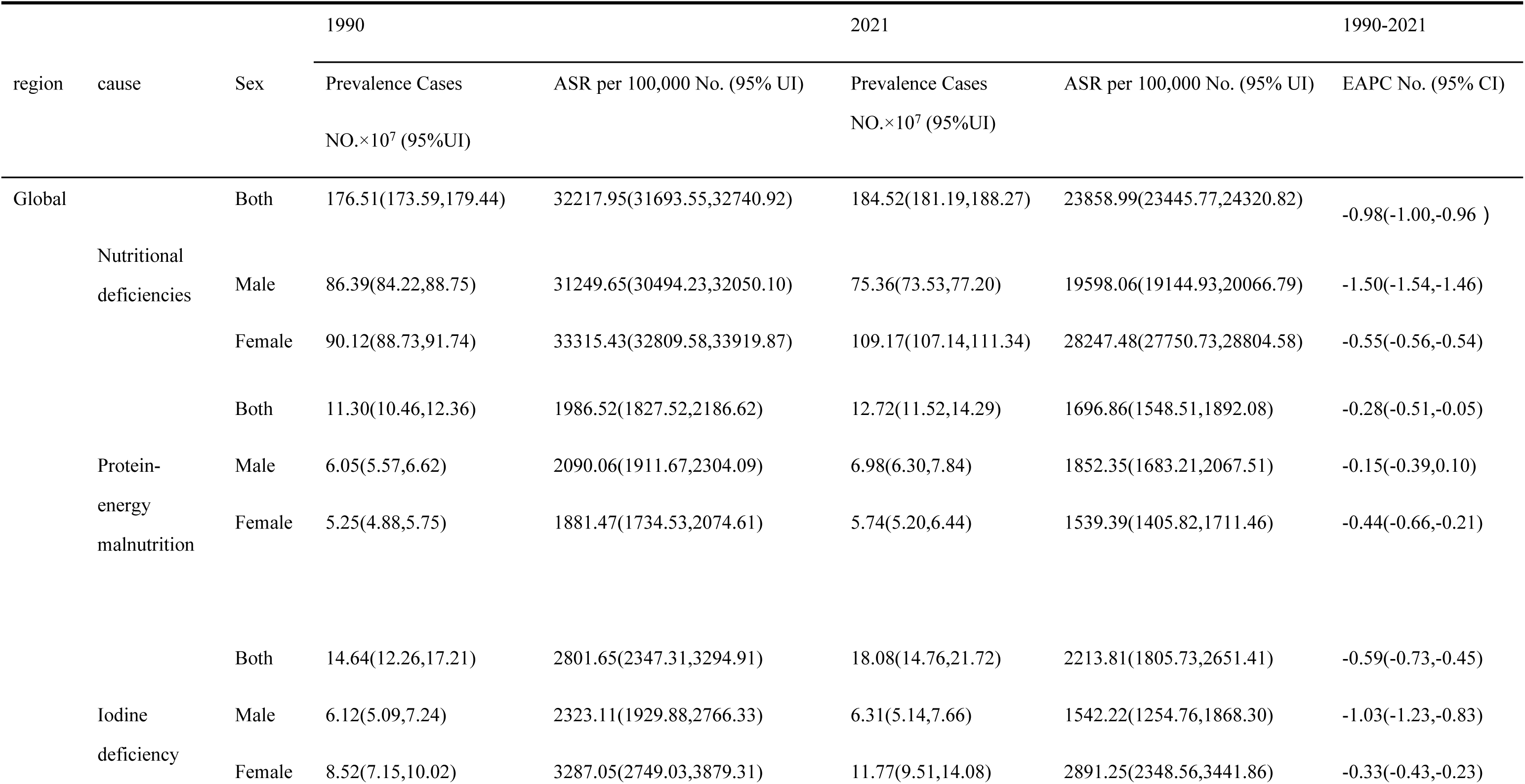
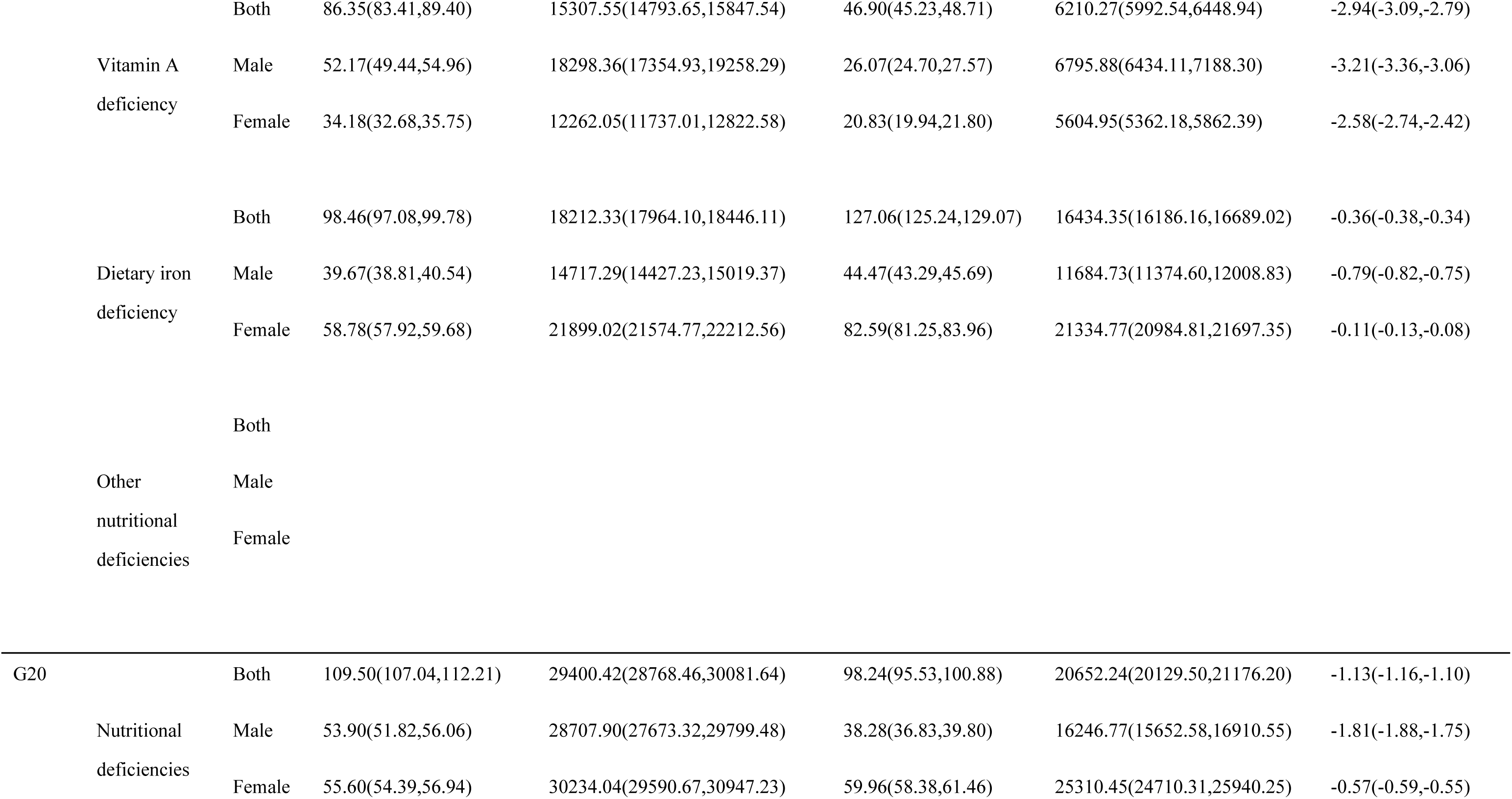
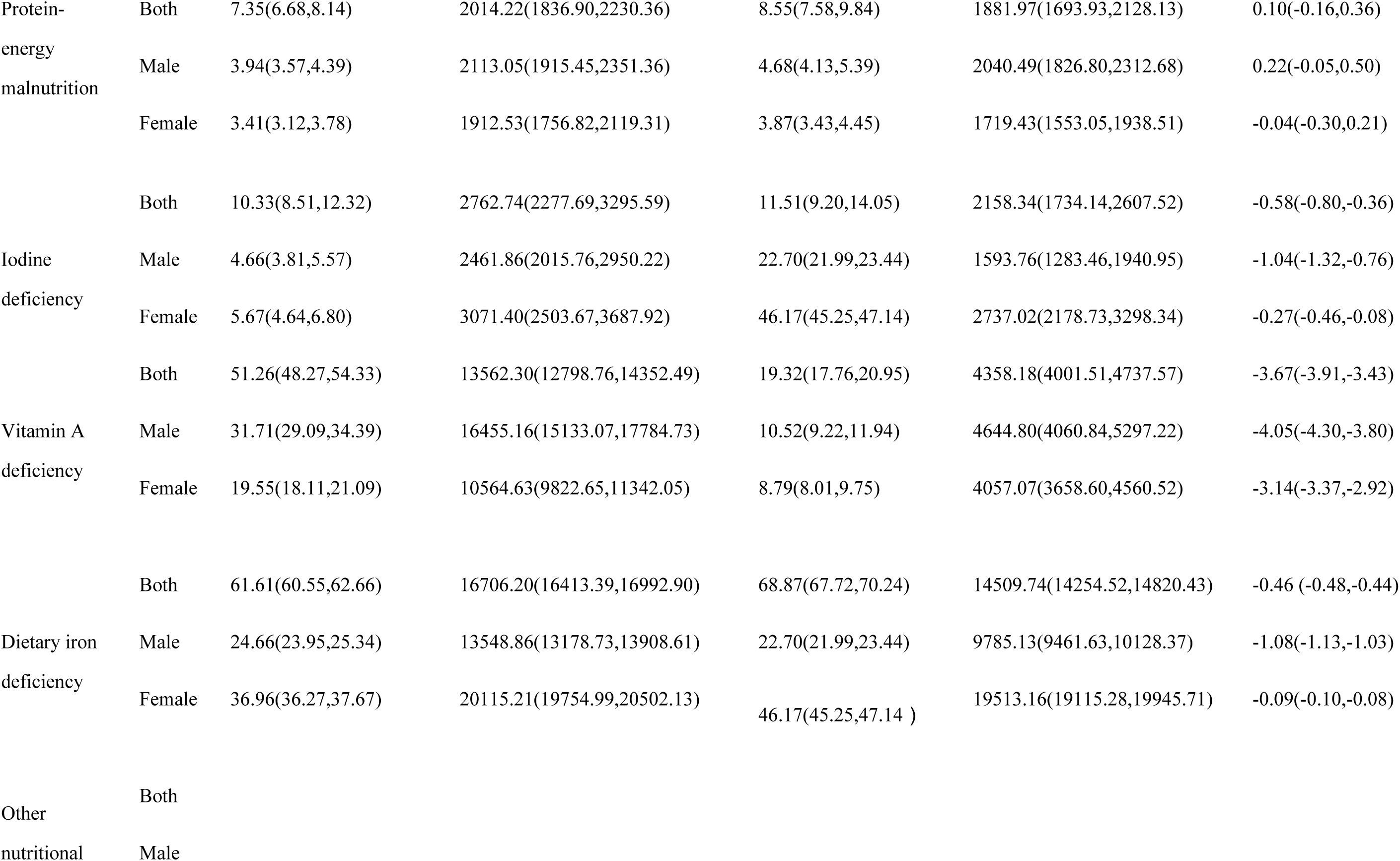
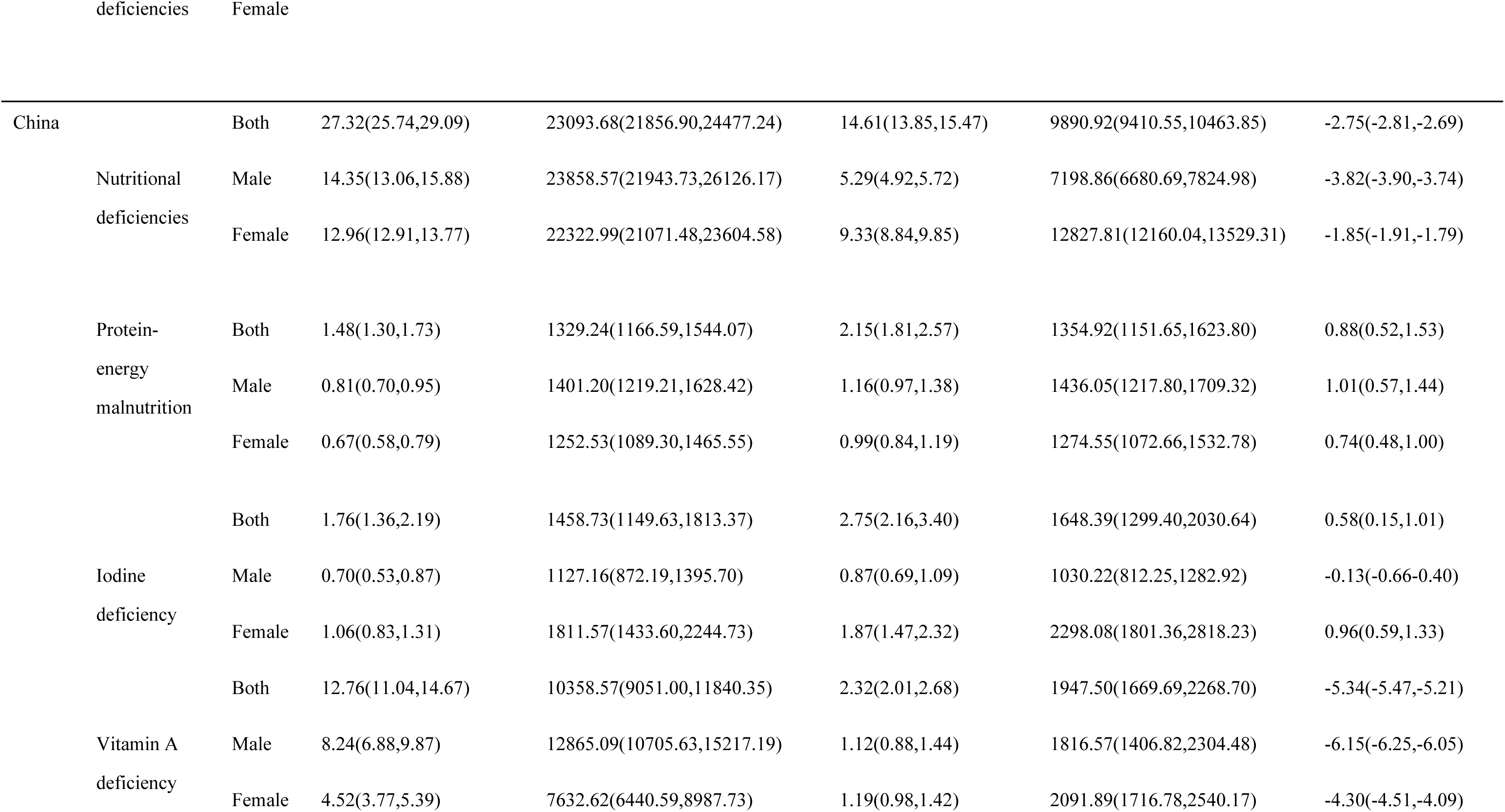
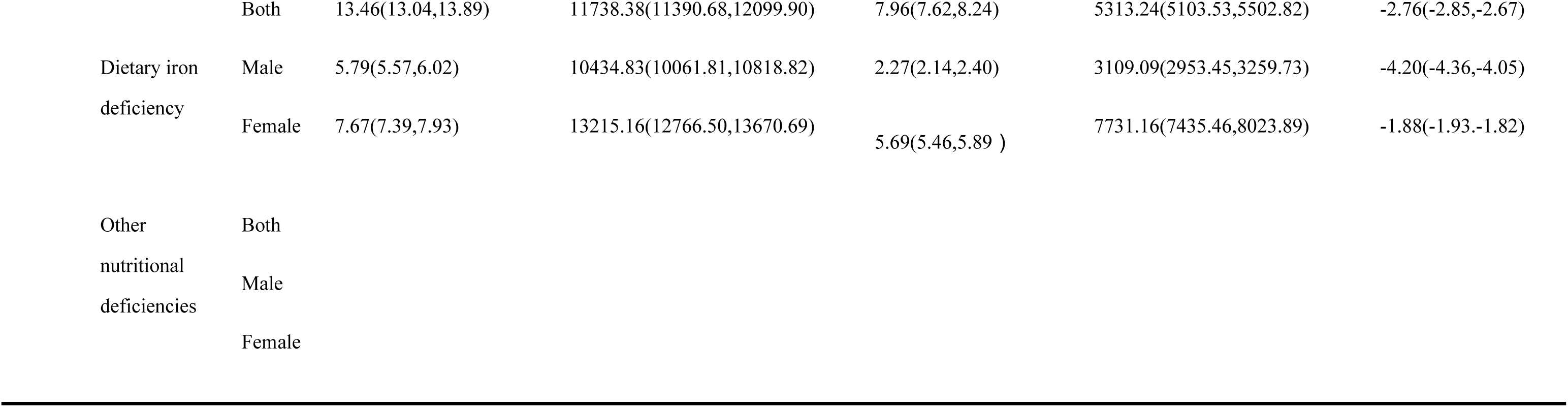
The prevalence levels of nutritional deficiency in China, the G20, and Globally from 1990 to 2021.

**Table 3.** The AAPC of AS YLDs rate and ASPR of NDs from 1990 to 2021.

| AAPC |  |  |  |
| --- | --- | --- | --- |
| Region | Sex | Prevalence | YLDs |
| Global | Both | -272.236(-275.398, -269.074) | -5.126(-5.266, -4.985) |
|  | Male | -380.091(-384.234, -375.949) | -5.898(-6.056, -5.739) |
|  | Female | -166.027(-168.280, -163.775) | -4.090(-4.254, -3.926) |
| G20 | Both | -284.184(-288.389, -279.978) | -5.621(-5.799, -5.444) |
|  | Male | -406.293(-411.463, -401.122) | -6.390(-6.558, -6.221) |
|  | Female | -159.466(-162.455, -156.477) | -4.397(-4.547, -4.247) |
| China | Both | -428.495(-433.504, -423.486) | -6.180(-6.219, -6.142) |
|  | Male | -543.072(-548.740, -537.404) | -7.100(-7.126, -7.073) |
|  | Female | -304.736(-309.343, -300.129) | -5.280(-5.337, -5.222) |

The prevalence in the G20 group attributed to nutritional deficiency in 2021 was approximately 0.98 billion cases (95% UI0.95–1.00 billion) with an ASPR of 20,652.24 cases per 100,000 population (95% UI: 20,129.50–21,176.20), compared to 29,400.42 cases per 100,000 population (95% UI: 28,768.46–30,008.16) in 1990. EAPC exhibited an average annual decline of 1.13% (95% CI: –1.16% to –1.10%), indicating a decreasing trend.

The prevalence in China, which was attributed to nutritional deficiency in 2021, was approximately 0.15 billion (95% UI: 0.14–0.15 billion), and the ASPR was 9,890.92 per 100,000 population (95% UI: 9,410.55–10,463.85), compared to 23,093.68 per 100,000 population (95% UI: 21,856.90–24,477.24) in 1990. The EAPC exhibited an average annual decrease of 2.75% (95% CI: –2.81% to –2.69%).

### 3.2 Disease burden of ND from 1990 to 2021 according to age and gender

Figure 1 illustrates that the highest incidence of the cases globally affects children < 5 years old, and male patients outnumber female patients. In the 5–9 years, male patients outnumber female patients. In all other age groups, female patients outnumber male patients.

**Figure 1.**
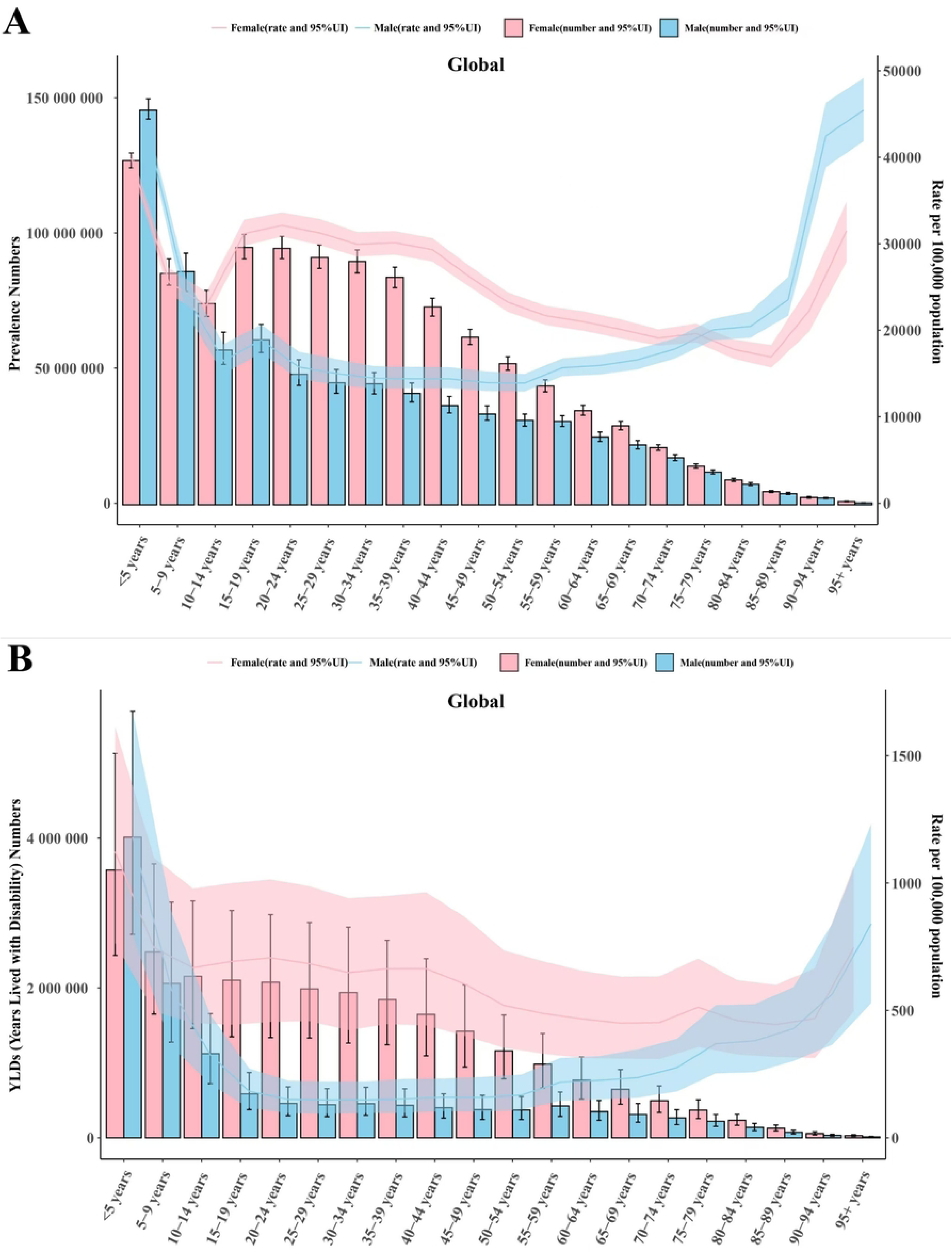
Age-standardized prevalence and YLDs rates for nutritional deficiencies in global from 1990 to 2021.

In the G20 group, the prevalence of malnutrition among children < 5 years old was the highest, and males outnumbered females. In other age groups, the number of female patients was generally higher than that of male patients. In China, individuals aged 30–34 had the highest prevalence of malnutrition, with female patients significantly outnumbering their male counterparts in this age group. Among those < 5 years old, males suffer more from malnutrition than females (Figure 2).

**Figure 2.**
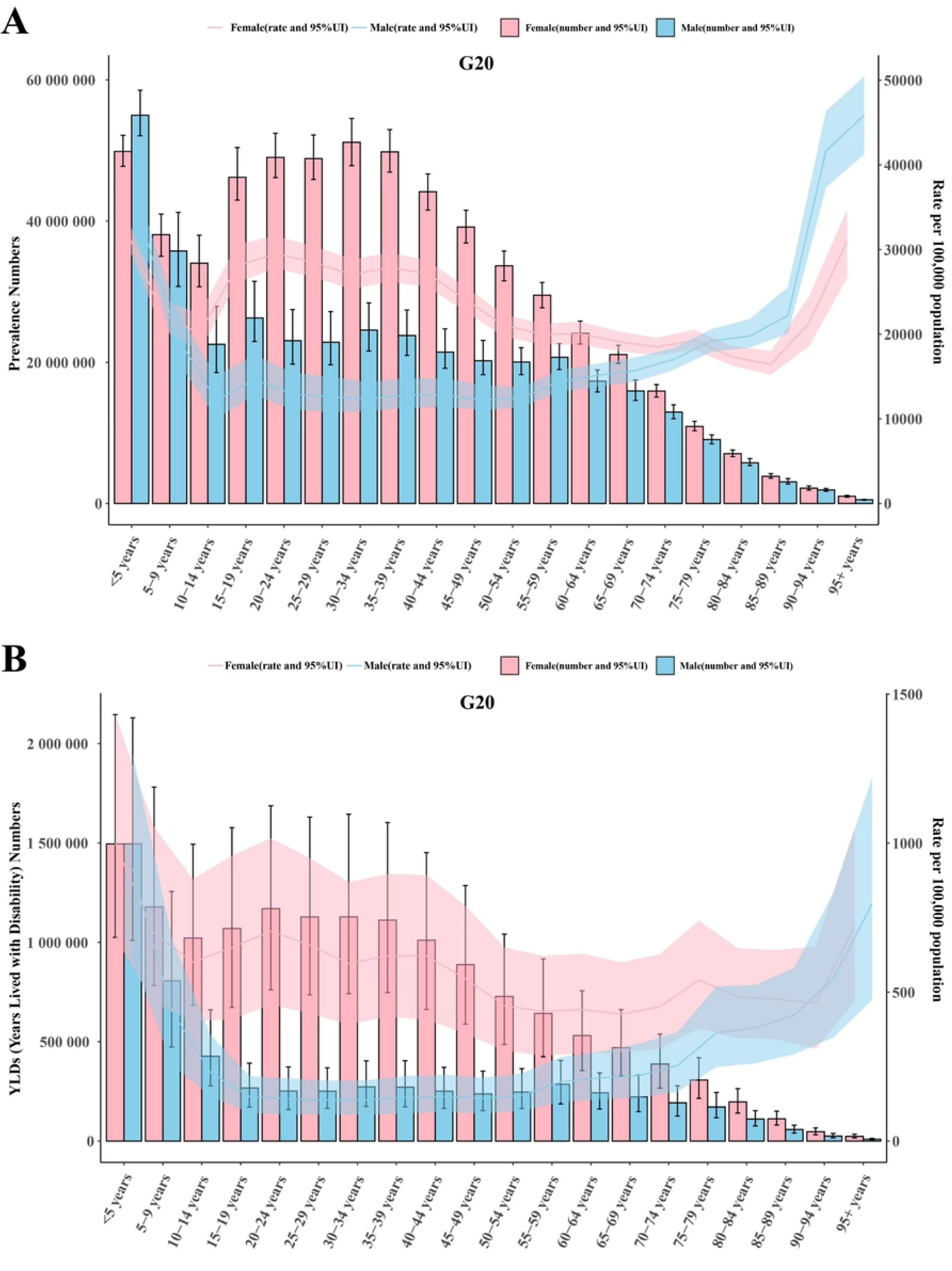
Age-standardized prevalence and YLDs rates for nutritional deficiencies in G20 group from 1990 to 2021.

Regarding the number of YLDs, the global population aged ≤ 5 had the highest number of YLDs due to malnutrition, with males outnumbering females. The situation was similar in the G20 group, where the number of YLDs attributed to malnutrition in the population aged ≤ 5 was highest, with males outnumbering females. In China, the number of YLDs attributed to malnutrition in the age group of 35–39 years was the highest, and the number of females attributed to malnutrition YLDs was the highest and significantly higher than that of males in the same age group. For children < 5 years old, the number of males attributed to malnutrition YLDs was higher than that of females.

In 2021, the global prevalence of malnutrition among individuals < 5 years old was the highest (Figure 1), while in China, it predominantly affected those aged between 20 and 60. Globally, in the G20 group and China, the prevalence of malnutrition among female patients exceeded that of male patients, particularly during pregnancy (Figure 3).

**Figure 3.**
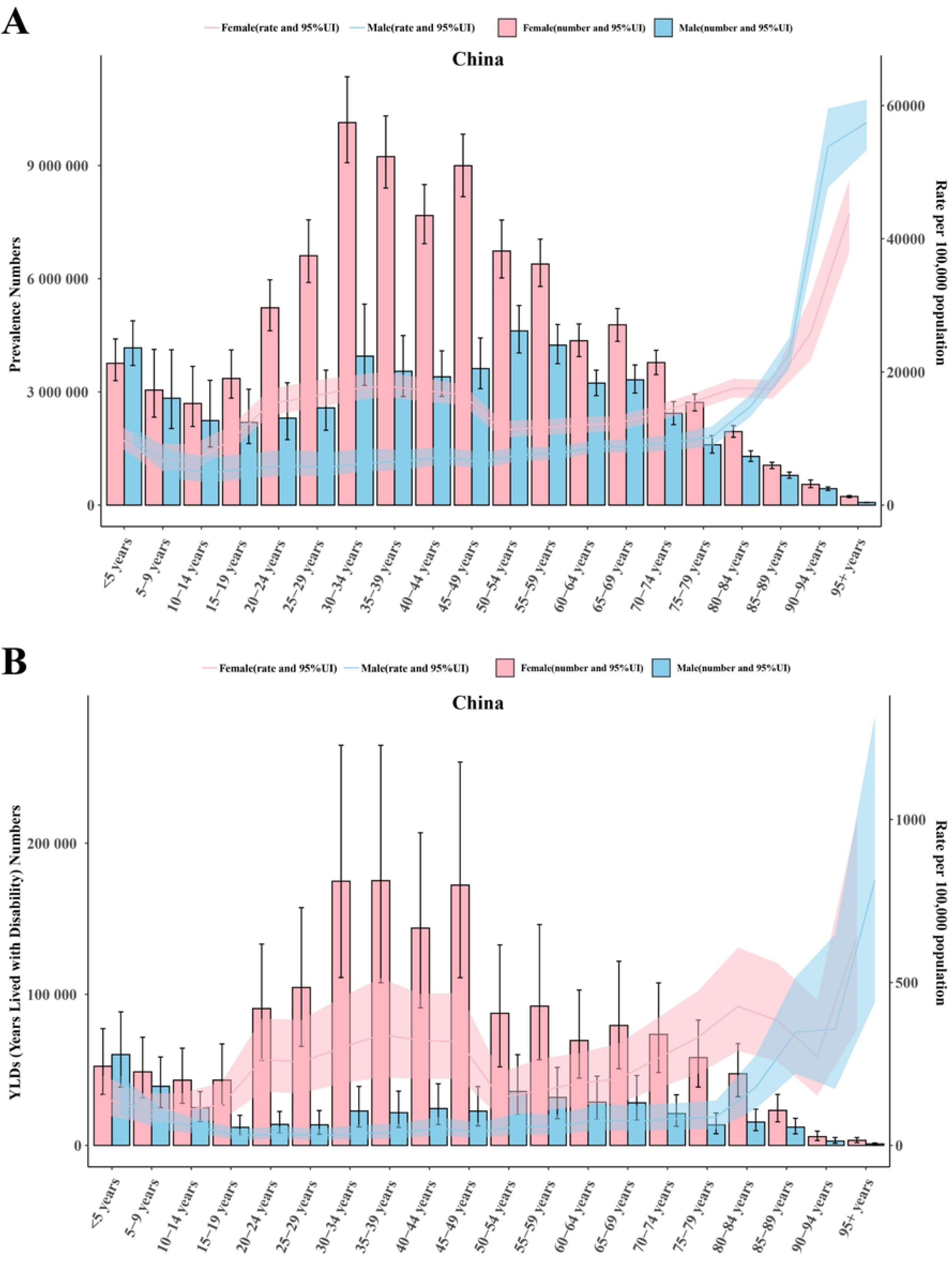
Age-standardized prevalence and YLDs rates for nutritional deficiencies in China from 1990 to 2021.

### 3.3 Analysis of disease differences

Tables 1 and 2 and Figure 4 indicate that among the subgroups of nutritional deficiencies, the global prevalence of dietary iron deficiency was highest in 1990 and 2021, with a higher proportion observed in females than in males. In the G20 group, the prevalence of dietary iron deficiency was highest, with a higher incidence observed in females than in males. This is true in China. Regarding prevalence, the global regional prevalence of dietary iron deficiency was highest in 1990 and 2021, with females exhibiting a higher prevalence than males. However, vitamin A deficiency was more prevalent in males than in females. Dietary iron deficiency is the most common nutritional deficiency in the G20 group, with a higher prevalence in females than in males. This is followed by vitamin A deficiency, which is more prevalent in males than in females. In China, dietary iron deficiency was the highest among nutritional deficiencies, with a higher prevalence in females than in males, followed by vitamin A deficiency, which is more prevalent in males than in females. Among the three regions, global, G20, and China, the prevalence of iodine deficiency was higher in females than in males, while the prevalence of PEM was higher in males than in females

**Figure 4.**
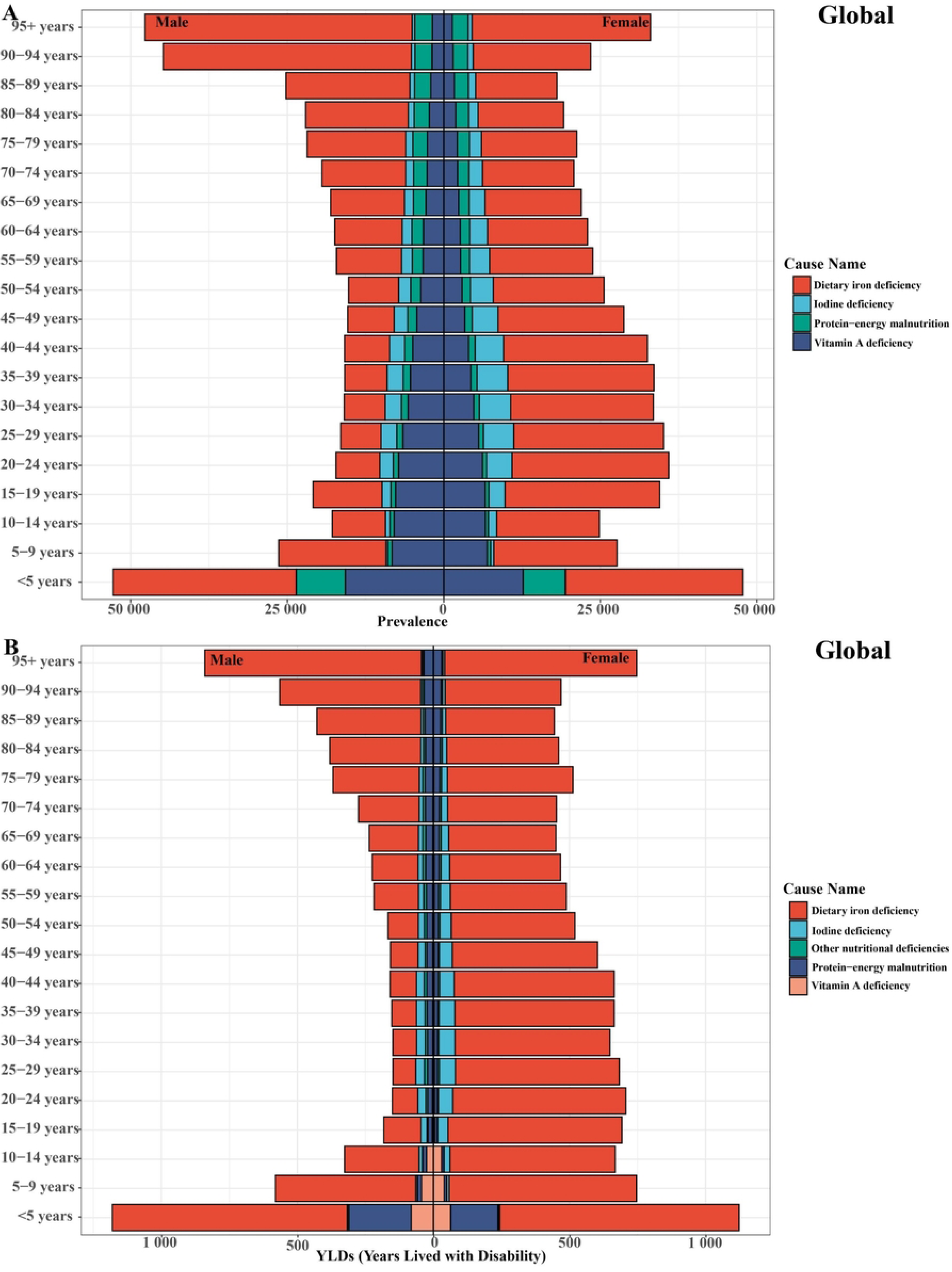
The analysis of disease differences for nutritional deficiencies in global from 1990 to 2021.

### 3.4 Joinpoint regression analysis and AAPC of the disease burden of ND from 1990 to 2021

AAPC is primarily used to describe the average annual change rate of a disease indicator (including incidence and mortality) over some time. It provides a more comprehensive trend analysis by integrating data from multiple years rather than simply comparing the data of the first two years.

We analyzed the age-standardized YLD rates and ASPR for the global, G20, and China from 1990 to 2021 (Figure 7). For the global age-standardized YLDs rate, the AAPC from 1990 to 2021 was 5.126 (95% CI: 5.266–4.985, P < 0.05). This period was divided into four parts at the connection points of 1995, 2004, and 2015. The age-standardized YLDs rate exhibited a declining trend in all four stages, with the fastest decline occurring from 2015 to 2021 APC: –1.339 (95% CI: –1.496 to –1.181, P < 0.05). The second fastest decline occurred from 2004 to 2015 APC: –1.122 (95% CI: –1.214, –1.030, P < 0.05). However, the decline was non-significant from 1995 to 2024 APC: –0.423 (95% CI: –0.525 to –0.340, P < 0.05), illustrating a relative stability. Regarding ASPR, the AAPC from 1990 to 2021 was –272.236 (95% CI: –275.398 to –269.074, P < 0.05). This period was divided into four stages at 1998, 2005, and 2008 connection points. ASPR exhibited a declining trend in all four stages, with the fastest decline occurring from 2005 to 2008 APC: –1.318 (95% CI: –1.652 to –0.984, P < 0.05). The second fastest decline occurred from 2008 to 2021 APC: –1.016 (95% CI: –1.052 to –0.981, P < 0.05). The least significant decline occurred from 1998 to 2005 APC: –0.806 (95% CI: –0.879, –0.733, P < 0.05).

**Figure 5.**
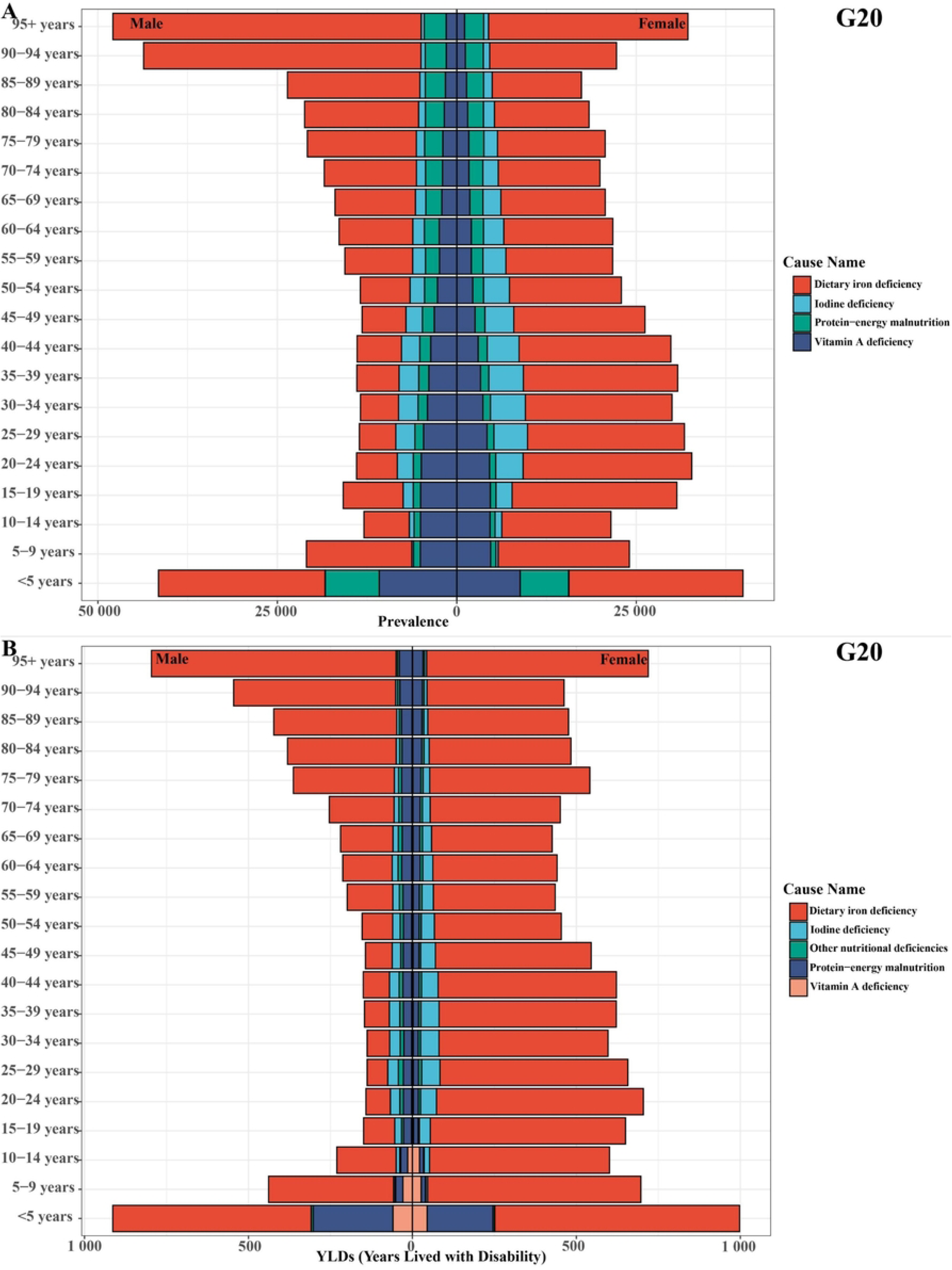
The analysis of disease differences for nutritional deficiencies in G20 group from 1990 to 2021.

**Figure 6.**
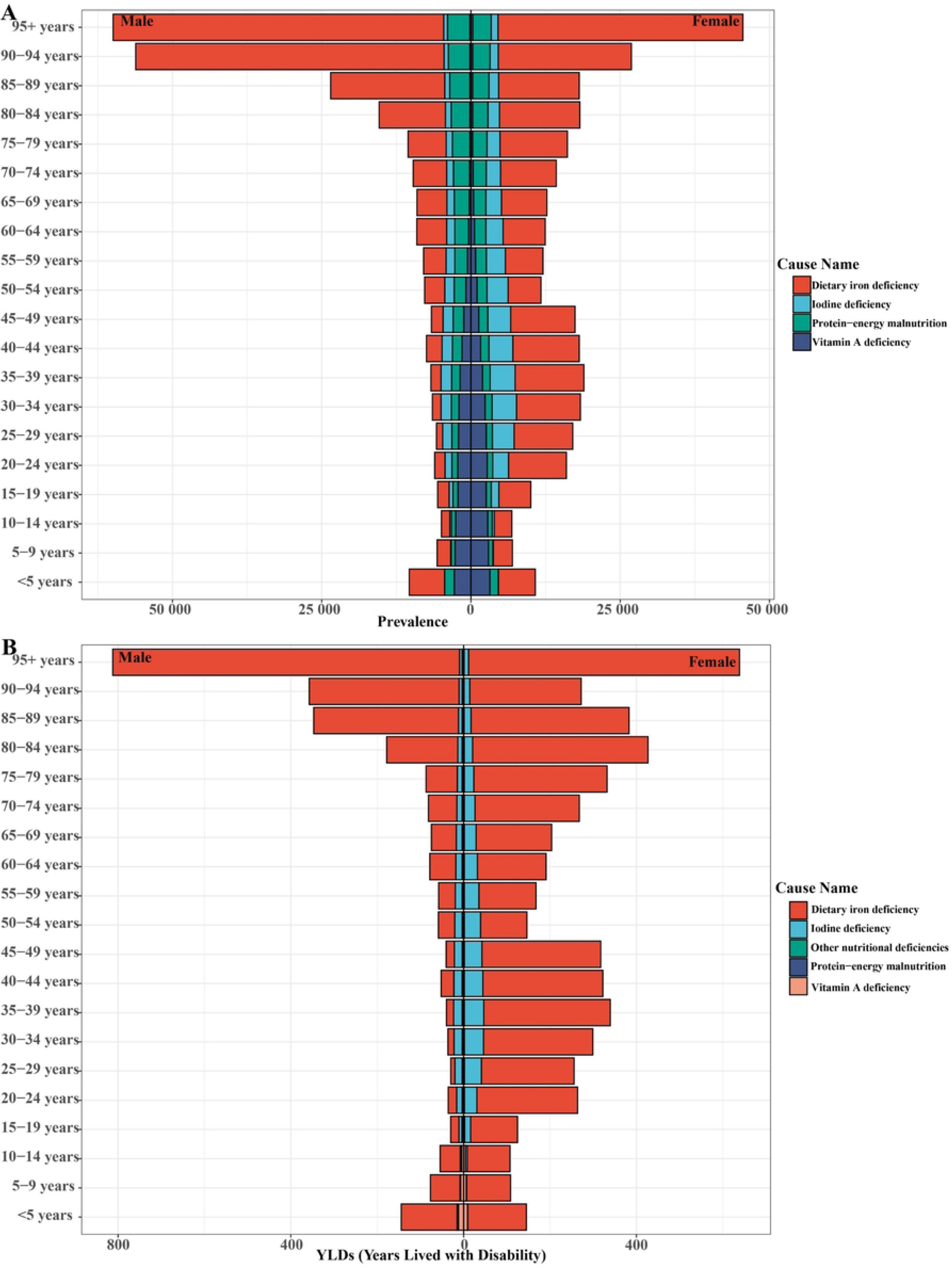
The analysis of disease differences for nutritional deficiencies in global from 1990 to 2021.

**Figure 7.**
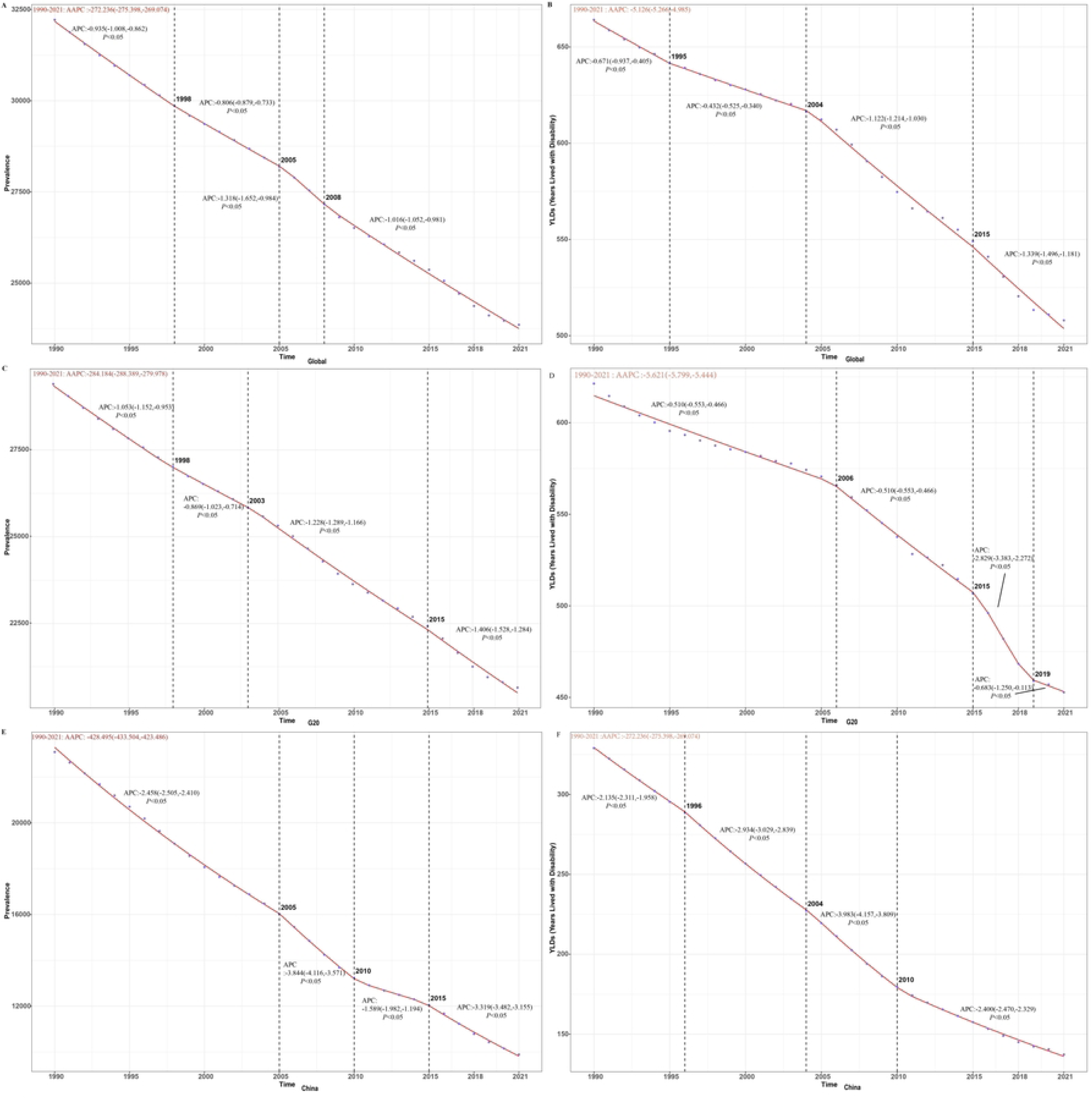
Joinpoint regression analysis and AAPC of the disease burden of ND in China, the G20, and Globally from 1990 to 2021.

The age-standardized YLD rate of nutritional deficiency in the G20 group from 1990 to 2021 indicated that the AAPC was –5.621 (95% CI: –5.779 to –5.444, P < 0.05). This period was divided into four stages at three connection points: 2006, 2015, and 2019. All four stages exhibited a declining trend. Among them, the fastest decline occurred from 2015 to 2019 APC: –2.829 (95% CI: –3.383 to –2.272, P < 0.05). The second fastest decline was from 2006 to 2015 APC: –1.190 (95% CI: –1.278 to –1.102, P < 0.05). The most non-significant decline occurred from 1990 to 2006 APC: –0.510 (95% CI: –0.553 to –0.466, P < 0.05). The AAPC for ASPR from 1990 to 2006 was –284.184 (95% CI: –288.389 to –279.978, P < 0.05). This period was divided into four stages at three connection points: 1998, 2003, and 2015. All four stages exhibited a declining trend. The fastest decline occurred from 2015 to 2021 APC: 1.406 (95% CI: –1.528 to –1.284, P < 0.05). The second fastest decline was from 2003 to 2015 APC: –1.228 (95% CI: –1.289 to –1.166, P < 0.05). The most non-significant decline occurred from 1998 to 2003 APC: –0.869 (95% CI: –1.023 to –0.714, P < 0.05).

The age-standardized YLD rate of Chinese nutritional deficiencies indicated that the AAPC from 1990 to 2021 was –6.180 (95% CI: –6.219 to –6.142, P < 0.05). This period was divided into four stages at three connection points in 1996, 2004, and 2010. All four stages exhibited a declining trend. The most significant decline occurred during the period from 2004 to 2010 APC: –3.983 (95% CI: –4.157 to 3.809, P < 0.05). The second most significant decline occurred from 1996 to 2004 APC: –2.934 (95% CI: –3.029, –2.839, P < 0.05). The period from 1990 to 1996 APC: –2.934 (95% CI: –3.029, –2.839, P < 0.05) exhibited the slowest decline, indicating stability. The AAPC from 1990 to 2021 was –428.495 (95% CI: –433.504 to –423.486, P < 0.05). This period was divided into four stages at three connection points in 2005, 2010, and 2015. All four stages exhibited a declining trend. The most significant decline occurred from 2005 to 2010 APC: –3.844 (95% CI: –4.116 to –3.571, P < 0.05). The second most significant decline occurred from 2015 to 2021 APC: –3.319 (95% CI: –3.482 to –3.155, P < 0.05). The most non-significant decline occurred from 2010 to 2015 APC: –1.589 (95% CI: –1.982 to –1.194, P < 0.05).

### 3.5 Decomposition analysis

This study analyzes the three primary factors contributing to nutritional deficiency: aging, population growth, and epidemiological changes (Figure 8). Between 1990 and 2021, population growth has been the primary factor contributing to the increased prevalence and YLD rates of global nutritional deficiency. Epidemiological shifts have primarily driven increased prevalence and YLD rates of nutritional deficiency in China. The impact of population growth on the global increase in YLD malnutrition rates was 733.51%. Epidemiological changes account for a 143.36% increase in the YLD rate of malnutrition in China. Population growth accounts for 819.92% of global malnutrition prevalence, while epidemiological changes contribute 109.04% to the overall increase in the prevalence in China.

**Figure 8.**
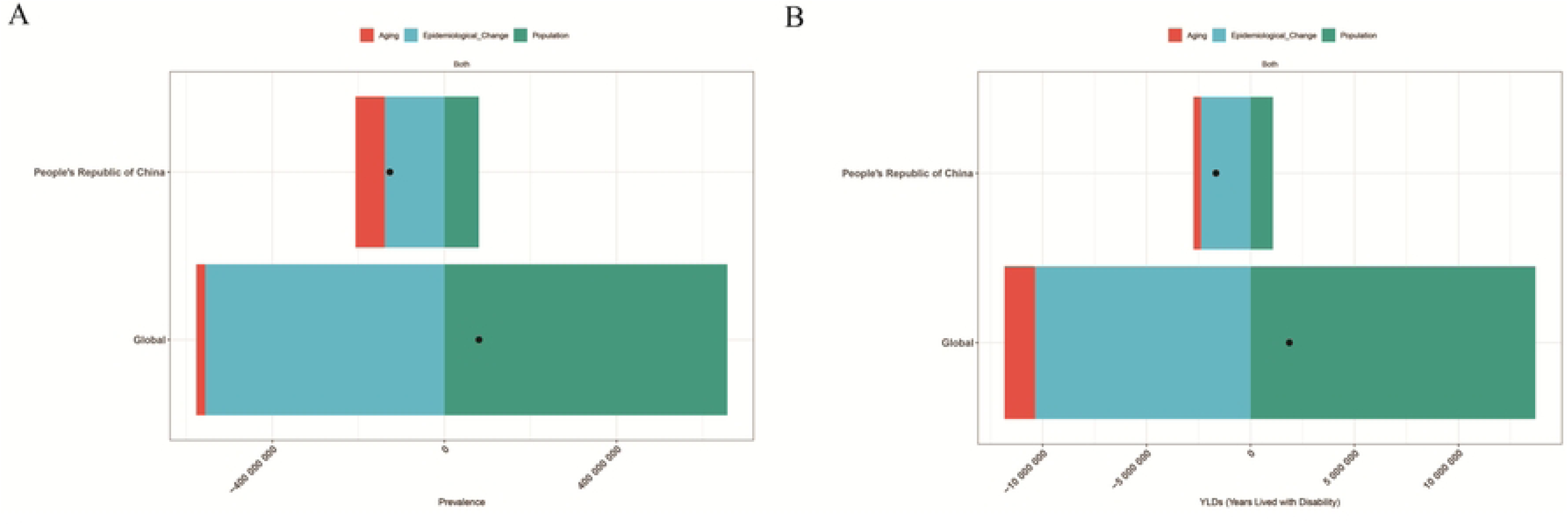
Decomposition of the number of cases of prevalent in global and China from 1990 to 2021.

### 3.6 Bayesian Age-Period Cohort model (BAPC)

The BAPC provides a comprehensive framework for executing full Bayesian inference using integrated nested Laplace approximations (INLA). BAPC is especially effective for predicting future ratios based on historical data, making it a valuable tool for public health planning and analysis.

Figure 9 illustrates our projections of the ASPR and the age-standardized YLD rate of nutritional deficiency in China and globally. Regarding gender, the global age-standardized YLD rate of nutritional deficiency is expected to decrease until 2035, with China exhibiting a consistent declining trend. The global ASPR of nutritional deficiency exhibited a declining trend, with China being no exception. In the 75–84 years age group, the decline in the ASPR of nutritional deficiency was most significant globally. The age-standardized YLD rate for nutritional deficiency decreases most significantly in the 80–89 years. The situation in China differs; the ASPR of nutritional deficiency decreases most significantly in the 85–89 years age group, while the age-standardized YLD rate decreases most significantly in the 25–39 years age group

**Figure 9.**
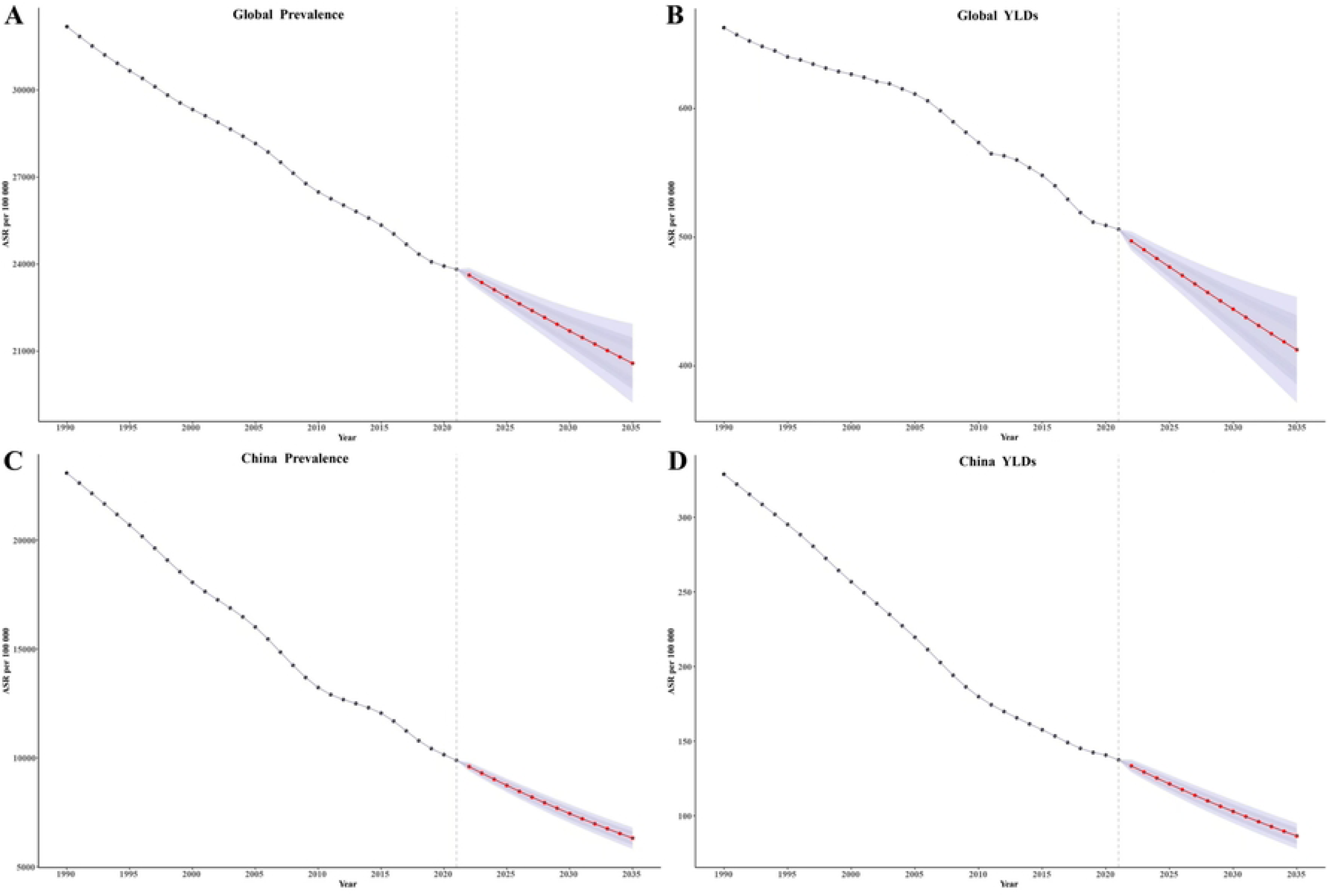
The BAPC model prediction for nutritional deficiency in China and global until 2035.

## 4. Discussion

The overall burden of nutritional deficiency in China From 1990 to 2021 exhibited a declining trend. Simultaneously, the prevalence of nutritional deficiency in the global and G20 regions exhibited a significant decline. In previous research spanning two decades from 1990 to 2019, the overall age-standardized incidence of nutritional deficiencies in China remained constant, while the overall age-standardized DALY rate declined. In 2019, the most severe nutritional deficiency subtypes were vitamin A deficiency, PEM, and dietary iron deficiency^[29]^. This study demonstrated that dietary iron deficiency exhibited the most significant disease burden among various subtypes of nutritional deficiencies. This may pertain to socioeconomic development in China. The economic development in China has been uneven due to geographical and other factors, leading to imbalances between the Eastern and Western regions and resulting in disparate levels of socioeconomic development. Prevention and treatment of dietary iron deficiency are the primary strategies to reduce the disease burden. Dietary intake of iron-rich foods or products is considered a safer remedy for iron deficiency than oral iron supplements^[29]^. Anemia caused by iron deficiency is the most common iron-related disease in China. Due to diverse dietary habits across different regions in China, a uniform dietary pattern affects the bioavailability of dietary iron, necessitating customization to align with regional dietary habits. Studies on dietary patterns are conducted to develop local recipes and improve the utilization of dietary iron.

In response to such circumstances, we can implement preventive measures from various perspectives, including re-optimizing the food structure, resolving supply-demand discrepancies, ensuring food availability, guaranteeing food safety and quality from production to consumption, and enhancing public well-being. Additionally, we need to popularize necessary nutritional knowledge and guide food consumption, emphasizing making the public understand balanced diet principles, the correlation between nutrition and health, and the connection between nutrition and disease, thereby improving the nutritional health awareness of the people. Prevention should be strategic, based on research and investigation, implementing specific and targeted preventive measures for diverse nutritional issues in different regions and populations.

These subtypes of nutritional deficiencies primarily affect children < 5 years old, potentially associated with their physical condition and nutritional status. Despite governmental efforts to enhance corresponding measures to improve the nutritional conditions of children, it is estimated that the global prevalence of anemia among children < 5 years old is 43%, with at least 50% affected by iron-deficiency anemia, one of the most common public health issues globally. Moreover, trace elements, including iron, iodine, and vitamin A, are crucial for the growth and development of children. Inadequate nutritional support for the growth and development of children can lead to deficiencies in these trace elements, resulting in unpredictable and irreversible damage. This adversely affects physical growth and psychological development, increasing the risk of diseases, infections, and mortality in children and subsequently impacting social productivity.

Furthermore, women experience a greater disease burden, especially those in the 30–34 age group. Unequal socioeconomic development, coupled with pressures experienced by women and the neglect of their nutritional status, results in numerous health issues. Regarding health, poverty and unequal socioeconomic development exacerbate the mental health issues and disease risks among women, resulting in malnutrition and adversely affecting their physical health. Economically, women face limited job opportunities and higher dependence on family economic support. Socially and culturally, poverty and unequal social development result in inadequate educational opportunities for women, lower social participation, and an increase in gender-based violence and discrimination against women. Regarding reproductive health, poverty and social inequality exacerbate fertility risks and sexual health issues for women. In populations with high iron requirements, including children, breastfeeding women, and adolescents, dietary iron deficiency can cause significant harm, resulting in significant economic losses.

Economic poverty and social inequality are social factors that may contribute to these disparities. The interaction between poverty and limited access to community healthcare can worsen this social inequality for women, resulting in a diminished likelihood of receiving timely treatment for their health conditions. Consequently, they may experience disabilities for longer periods than men^[28]^. This study has some limitations. First, computational resources preclude the propagation of uncertainty in specific covariates throughout the analysis, encompassing model estimates and life tables at each stage and uncertainties in key demographic inputs and outputs while excluding some covariates, including lagged income distribution and education. Similar limitations apply to other covariates used in subsequent analytical steps, including the completeness of age groups > 5 years old in the comprehensive completeness model, which does not account for measurement errors^[29]^.

Second, the original data obtained from this study was limited, and all came from GBD 2021. Consequently, the findings of this study largely depend on the data from GBD 2021. In practice, accessing key data may be infeasible, or obtaining such data may be difficult^[30]^. Therefore, ensuring the quality of data collection in national health systems is crucial, and public health professionals can evaluate the efficacy of local guidelines and policies through comprehensive analysis of these data. Cohort and prospective studies are essential for investigating health concerns across various populations. Epidemiology employs prospective studies to examine disease trends within populations. Simultaneously, epidemiology can be combined with psychology, genetics, and etiological research.

The APC model employed in this study is mathematically derived and theoretically biased, rendering it incapable of fully representing real-world situations; accordingly, the APC model should be utilized judiciously when interpreting the results^[31]^. The existing theoretical framework of this study has certain limitations and cannot comprehensively explain or cover all phenomena and issues encountered in the research.

## 5. Conclusion

The disease burden of nutritional deficiency in China demonstrated a declining trend from 1990 to 2021, which may be associated with the socioeconomic development of China. The prevalence of dietary iron deficiency-related diseases may correlate with variations in dietary patterns across regions in China. The disease burden is greater among children < 5 years old, associated with their nutritional status, necessitating the implementation of appropriate measures to improve the nutritional status of this population to reduce the disease burden. Nutritional deficiencies are critical indicators of malnutrition, and eliminating the damage caused by malnutrition is difficult

## Data Availability

yes

## Acknowledgments

Our gratitude goes to the participants of this research for their collaborative efforts. Our gratitude extends to the School of Public Health at Shaanxi University of Chinese Medicine for their assistance.

## Author contributions

LM holds the responsibility for data visualization and writing;

XL and XZ contributed to writing-review and editing;

RZ and LM oversees certain aspects of data management;

RZ helped with supervision, performed project administration, and helped in funding acquisition.

All authors have read and agreed to the published version of the manuscript.

Informed consent

Informed consent was not needed.

## Ethics

The institutional review board granted an exemption for this study, as it utilized publicly accessible data that contained no confidential or personally identifiable patient information.

